# Human *in vivo* footprints from blood plasma samples for improved diagnostics in septic patients

**DOI:** 10.1101/2025.01.29.25320179

**Authors:** Mirko Sonntag, Jan Müller, Thorsten Brenner, Sebastian O. Decker, Arndt von Haeseler, Kai Sohn, the German Anaesthesia and Intensive Care Trials Group (GAIN-CARE)

**Affiliations:** Innovation Field In-Vitro Diagnostics, Fraunhofer Institute for Interfacial Engineering and Biotechnology IGB, 70569 Stuttgart, Germany; Interfaculty Graduate School of Infection Biology and Microbiology (IGIM), Eberhard Karls University Tuebingen, 72076 Tuebingen, Germany; Center for Integrative Bioinformatics Vienna, Max Perutz Labs, University of Vienna, Medical University of Vienna, 1030 Vienna, Austria; Vienna Biocenter PhD Program, a Doctoral School of the University of Vienna and the Medical University of Vienna, 1030 Vienna, Austria; Department of Anesthesiology and Intensive Care Medicine, University Hospital Essen, University Duisburg-Essen, 45147 Essen, Germany; Department of Anesthesiology, Medical Faculty Heidelberg, Heidelberg University, 69120 Heidelberg, Germany; Ludwig Boltzmann Institute for Network Medicine, University of Vienna, Augasse 2-6, 1090 Vienna, Austria; Heidelberg University, Medical Faculty Heidelberg, Department of Anesthesiology, Im Neuenheimer Feld 420, 69120 Heidelberg, Germany; Department of Anesthesiology and Intensive Care Medicine, University Hospital Essen. University Duisburg-Essen, Hufelandstr. 55, 45147 Essen, Germany; Fraunhofer IGB, Nobelstr. 12, 70569 Stuttgart, Germany; Institute of Medical Biometry and Informatics, University of Heidelberg, Im Neuenheimer Feld 130.3, 69120 Heidelberg, Germany; Coordination Centre for Clinical Trials (KKS), Ruprecht-Karls-University, Berliner Straße 10, 69120 Heidelberg, Germany; Center for Infectious Diseases and Infection Control, Jena University Hospital, Erlanger Allee 101, 07740 Jena, Germany; University Ulm, Faculty of Medicine, Department of Anaesthesiology & Intensive Care Medicine, Ulm, Germany; Technical University of Munich, School of Medicine and Health, Department of Anaesthesiology and Intensive Care Medicine, Munich, Germany; Abteilung Klinische Infektiologie, Evangelisches Klinikum Bethel, Universitätsklinikum OWL der Universität Bielefeld, Campus Bielefeld Bethel, Bielefeld, Germany; Department of Anesthesiology, University Medical Center Göttingen (UMG), Medical University of Göttingen, Göttingen, Germany; Department of Anesthesiology, Medical Faculty and University Hospital Duesseldorf, Heinrich-Heine-University Duesseldorf, 40225 Duesseldorf, Germany; Klinik für Anästhesiologie und Intensivmedizin, Medizinische Hochschule Hannover, Hannover, Deutschland; Anästhesie und Intensivmedizin, Evangelisches Krankenhaus Luckau, Luckau, Deutschland; Department of Anesthesiology and Intensive Care Medicine, University Hospital Tuebingen, Eberhard-Karls-University, Hoppe-Seyler-Strasse 3, 72076 Tuebingen, Germany; Department of Anesthesia and Intensive Care, Rostock University Medical Center, Rostock, Germany; Department of Intensive Care and Intermediate Care, University Hospital RWTH Aachen, Aachen, Germany; Goethe University Frankfurt, Department of Anaesthesiology, Intensive Care Medicine and Pain Therapy, University Hospital Frankfurt, 60590 Frankfurt, Germany; Charité - Universitätsmedizin Berlin, corporate Member of Freie Universität Berlin and Humboldt-Universität zu Berlin, Department of Anesthesiology and Intensive Care Medicine (CCM/CVK), Berlin, Germany; Department of Anesthesiology and Surgical Intensive Care Medicine, Division of Intensive Care Medicine, Universitätsklinikum Bonn, Bonn, Germany; Department of Anesthesiology and Intensive Care Medicine, Faculty of Medicine and University Hospital of Cologne, University of Cologne, Kerpener Straße 62, 50937 Cologne, Germany; Medical ICU, University Hospital Leipzig, Leipzig, Germany; Department of Anaesthesiology and Intensive Care Medicine, General Hospital of Heidenheim, Heidenheim, Germany; Department of Nephrology, Heidelberg University Hospital, Heidelberg, Im Neuenheimer Feld 162, Germany

**Keywords:** Next-Generation diagnostics, Short cell-free DNA, Biomarker identification, Sepsis, Liquid biopsy

## Abstract

**Background:** Liquid biopsy based on cell-free DNA (cfDNA) is an established approach in clinical diagnostics. In recent years, a fraction of cfDNA comprising short fragments has been discovered, that is enriched at gene promoters and binding sites of DNA-binding proteins. However, the diagnostic potential of such short double-stranded cell-free DNA (footprint DNA) remains to be fully explored. Therefore, we characterized the clinical utility of footprint DNA in septic patients.

**Methods:** We enriched for footprint DNA based on size selection and subsequent high-throughput DNA sequencing to receive an unbiased, genome-wide picture of the host response to the infection. Footprint DNA occupancies were analyzed for correlation with clinical metrics including urea, hemoglobin, or alanine aminotransferase (ALT). Additionally, footprint DNA markers were benchmarked by read and receiver operating curve (ROC) analysis against procalcitonin (PCT) as an established marker for infection status as well as against clinical parameters for early death prediction.

**Findings:** We found that levels of occupancy of footprint DNA at defined genomic loci semi-quantitatively correlated with physiological markers like ALT or urea from major organ systems including liver or kidney. In a small proof-of-concept cohort, differential signatures of DNA footprints distinguished between patient groups with bacterial and viral infections with an area under the ROC (AUROC) of 1.0, which is considerably better than PCT with an AUROC of 0.75. Likewise, footprint DNA could also predict early death in septic patients with an AUROC of 0.983, compared to the SOFA (Sepsis-related organ failure assessment) score with an AUROC of 0.76.

**Interpretation:** Our findings show that footprint DNA delivers quantitative information on physiology at the DNA level, demonstrating its diagnostic and prognostic potential. Identified footprint biomarker regions could be helpful in the clinical assessment of septic patients and other complex diseases outperforming current state-of-the-art clinical diagnostics.

**Funding:** This study was financed with internal funds from Fraunhofer society.

## Introduction

The potential of liquid biopsy in clinical diagnostics has revolutionized the field of clinical diagnostics for multiple indications [1]. Especially cell-free DNA is widely used as a biomarker class for prognosis and diagnosis in non-invasive prenatal testing, cancer, sepsis and organ rejection, among others [2–4]. Exemplarily, several studies underline the potential of cfDNA based biomarkers for clinical sepsis diagnostics [5–7]. For pathogen detection in sepsis, it is well known that the clinical gold standard of culture-based analysis (e.g., blood culture) is associated with considerable limitations [8–10]. Using metagenomic high-throughput sequencing of plasma cfDNA, a six times higher sensitivity for pathogen detection than blood culture can be achieved [11]. More recently, in contrast to regular cfDNA, with an average fragment size of 167 bp, a very small subset of double-stranded cfDNA with a significantly shorter fragment length of 35 - 80 bp has been found to be protected by non-histone DNA-binding proteins, like transcription factors [12,13]. Using ultra-deep high-throughput DNA-sequencing, it has been described that short cfDNA directly maps to genomic regions that are occupied by DNA binding proteins like transcription factors, indicating first potential for clinical application for example in breast cancer diagnostics [12,14]. Alternative approaches to infer the transcription factor occupancy make use of nucleosomal positioning therefore detecting open chromatin, where DNA regulatory binding protein may bind [15]. Quite recently, we have developed a workflow to specifically enrich short double-stranded cfDNA from plasma (footprint DNA) and could show that these fragments accumulate at open chromatin and gene regulatory elements [16]. The signals extracted from the footprint DNA were found to have the clinical potential to distinguish between two different types of gastrointestinal cancer as well as to discriminate sepsis from non-infected postoperative controls and healthy individuals [16].

Sepsis, defined as a “life-threatening organ dysfunction caused by a dysregulated host response to infection” [17], is a major health threat with high mortality rates and a global incidence of estimated 48.9 mio sepsis cases with 11 mio deaths per year [18]. However, precise and reliable biomarkers for sepsis diagnosis and prognosis are still in their infancies. One standard assessment for sepsis diagnosis represents the Sequential (Sepsis-related) Organ Failure Assessment (SOFA) score, which evaluates multiorgan failure in ICU patients [19] and has been described to have predictive value for mortality risk [20,21]. Another important clinical parameter frequently used in sepsis is procalcitonin (PCT) which is an indicator for bacterial sepsis and primarily suggested to guide antibiotic treatment [22–24]. Still, PCT and SOFA, although routinely used in clinical practice, do not represent reliable predictors of infection and outcome, thus indicating that further research is needed to overcome limitations and to establish more precise biomarkers [25,26].

In this study, we evaluated the diagnostic potential of footprint DNA in the context of sepsis. Consequently, we identified footprint DNA target regions that have the potential to semi-quantitatively retrace patient physiology. Based on the host response, footprint DNA allowed to more accurately distinguish between bacterial and viral sepsis and to predict early deaths versus survivors better than established biomarkers, including SOFA and PCT.

## Methods

### Ethics Approval and patient consent

Septic patient samples from the monocentric clinical study S-097/2013, conducted in the surgical intensive care unit of Heidelberg University Hospital (November 2013 - January 2015, German Clinical Trials Register: DRKS00005463) [11] and from the multicentric study S-084/2017 (March 2019 - August 2020, German Clinical Trials Register: DRKS00011911, ClinicalTrials.gov: NCT03356249) [27] were used, with study and control patients or their legal representatives signing written informed consent. Both studies were conducted in accordance with the Declaration of Helsinki and the professional code of conduct for physicians of the competent state medical association in the current versions, approved by the Institutional Ethics Committee of the Medical Faculty of Heidelberg University. Samples from S-097/2013 were used for correlating clinical metrics with footprint DNA signals [11,28]. For infection type and outcome prediction, septic samples from the study S-084/2017 [27] and postoperative, non-infection controls from S-097/2013 [11,28] were used.

### Blood plasma preparation and cell-free DNA isolation

Plasma and cfDNA was isolated as described previously [11]. CfDNA was isolated with the QIAsymphony SP and the QIAsymphony DSP Circulating DNA Kit as well as the QiaCube connect and the QIAamp MinElute ccfDNA kit (Qiagen, Hilden, Germany) according to the manufacturer’s instructions. Eluted cfDNA was quantified with the Qubit dsDNA HS Assay Kit (Thermo Fisher, Waltham, USA) and cfDNA quality was assessed by the Fragment Analyzer High Sensitivity DNA Kit (Agilent, Santa Clara, California, USA).

### Library generation and Next-Generation sequencing

Footprint DNA sequencing libraries from isolated cfDNA were prepared as already described in [16], with 15 ng of cfDNA as input and final elution in 20 µL of nuclease-free water. Library generation with the NEXTFLEX Cell free DNA-Seq Kit (V2) (Perkin Elmer, Pittsburgh, USA) was automated using the Biomek FXP (Beckman Coulter, Brea, USA). Library quality was assessed with the Fragment Analyzer High Sensitivity DNA Kit (Agilent, Santa Clara, USA) and concentration was measured by the Qubit dsDNA HS Assay Kit (Thermo Fisher, Waltham, USA). Size selection of cfDNA libraries was performed as described in [16] using BluePippin and 3% agarose cassettes (Sage Science, Beverly, USA) with one minor modification: following the second size selection, samples were purified with 1.8 x the volume of AMPureXP beads (Beckman Coulter, Brea, USA). Size selection performance was evaluated by the Fragment Analyzer High Sensitivity DNA Kit (Agilent, Santa Clara, USA) and by Qubit dsDNA HS Assay Kit (Thermo Fisher, Waltham, USA). After size selection and molarity normalization, sequencing was performed with a NextSeq 2000 (Illumina, San Diego, USA) using 100 bp single read kits with a targeted sequencing depth of 50 million reads per sample.

### Raw data processing

After quality control of raw sequencing reads with FastQC (v0.12.1), the following steps were performed to remove sequencing artifacts: 1) Removal of sequencing adapters, terminal polyG sequences (min 10 G’s), and quality trimming (BBTools - bbduk.sh v39.01). 2) Removal of terminal single A introduced by library preparation (BBTools - bbduk.sh v39.01). 3) Size selection of sequenced reads allowing lengths greater than 20 bp and smaller than 60 bp (BBTools - bbduk.sh v39.01). 4) Removal of sequencing reads with dust scores smaller than 7 (prinseq-lite v0.20.4) [29–31]. 5) Processed reads were mapped to the human reference genome GRCh37 using NextGenMap (v0.5.5) with default settings [32]. 6) Mapped reads were deduplicated with samtools rmdup (v1.6), reads in blacklisted regions were removed, and reads with a MAPQ value < 1 were removed with samtools view (v1.6) [33,34]. Resulting reads were converted to BigWig for visualization and other downstream analyses with deeptools (bin size = 10, normalization = counts per million (CPM), bamCoverage v3.5.2) [35].

### Peak calling and annotation

For footprint DNA data, peaks were called with MACS2 callpeak (narrow: --nomodel --extsize 32 --call-summits --min-length 30 -q 0.05; v2.2.9.1) [36]. Consensus peaks of a condition were identified with R if narrow peaks were identified in at least fifty percent of samples of a given condition. In addition, consensus peaks less than 31 nucleotides apart were merged. Consensus peaks were annotated to genomic functional elements with bedtools (nuc; v2.30.0) [37] including gene bodies, CpG islands, cis-regulatory elements (CREs) and transcription factor binding sites (TFBS). References for functional elements are provided on github and were originally retrieved from UCSC (CpG islands: UCSC TableBrowser, track name=cpgIslandExt; TFBS: UCSC TableBrowser, track name=encRegTfbsClustered) and ENCODE (CREs: https://doi.org/doi:10.17989%2FENCSR461KLY, https://doi.org/doi:10.17989%2FENCSR471KRT, https://doi.org/doi:10.17989%2FENCSR676GWZ, https://doi.org/doi:10.17989%2FENCSR405FRQ).

### Transcription factor motif enrichment analysis

Enriched transcription factor motifs were identified with MEME (AME; --scoring avg -- method fisher; v5.4.1) [38]. Input DNA sequences were retrieved from consensus peaks with bedtools (nuc; v2.30.0) and analyzed for enrichment of motifs in the HOCOMOCOv11_core_HUMAN_mono database [37,39]. DNA sequences from each condition’s consensus peak set served as test or control, e.g., consensus peak DNA sequences from bacterial sepsis as test for consensus peak DNA sequences from viral sepsis as control. Motifs with adjusted p-values < 0.05 were considered significantly enriched.

### Identification of differentially occupied genomic regions

Differentially enriched genomic regions were identified from the respective consensus peak sets of two conditions of sepsis outcomes or sepsis infection types. Genomic GC content of the consensus peak set used in a comparison was extracted with bedtools (nuc; v2.30.0), for later normalization of read counts [37]. Raw read counts were obtained with deeptools (multiBamSummary; v3.5.2) [35]. The read count matrix, GC content information and experiment design were used as input for differential enrichment analysis in R with EDASeq and edgeR packages (EDASeq v2.32.0; withinLaneNormalization (y=’GC’, which=’full’), betweenLaneNormalization (which=’full’), glmFit, glmLRT) [40,41]. Genomic regions with an adjusted p-value < 0.05 and a |log2(fold change)| > 0.5 were considered as differentially enriched regions (DERs). In addition, regions were ranked by p-values per fold change direction. From the ranked regions, the top 5 per fold change direction were selected. The top 5 DERs for the early death condition of the outcome comparison also includes regions with an adjusted p-value > 0.05. Normalized read counts from top regions were visualized as heatmaps with gplots (heatmap.2; v3.1.3; log2(normalized read count + 1), z-scaling per region, clustering by regions and samples using Euclidean distance and ward.D2 clustering). Normalized read counts of the top regions per condition were used as input for a principal component analysis (PCA) in R (prcomp; center=TRUE, .scale=TRUE). The first two principal components were evaluated for their discriminative power of the experimental conditions with receiver operating characteristic (ROC) analysis in R (pROC; v1.18.0).

For the infection type comparison, bacterial and viral sepsis samples were separately compared to non-infected control samples. All three comparisons, bacterial vs. viral, bacterial vs. no infection, and viral vs. no infection, yielded each 10 top DERs, in total 30. The GC content, sequencing depth, and region size normalized read counts from these regions were used for PCA.

### Correlation analyses between clinical metrics and short cfDNA

For the correlations between footprint DNA and clinical metrics, two patient groups were used. The ’Identification’ cohort (n = 28) included time course samples from six sepsis patients over 21 days, with eight samples missing due to unavailability or exclusion as described earlier [11]. Afterwards, identified correlates were validated with independent samples of varying time points from additional sepsis patients (‘Test’ cohort, n = 34). Correlations were derived from read counts of genomic bins to ensure an analysis that encompasses the entire genome, rather than focusing solely on regions with detectable peaks in the samples. This approach allows the detection of variations across the full dynamic range of read counts, including low or zero values, which might be missed by peak-calling methods. The genome was split in 30,962,143 bins of length 100nt and the short cfDNA reads per bin were counted. Read counts of bins were adjusted for GC content in R with EDASeq (EDASeq v2.32.0; withinLaneNormalization (y=’GC’, which=’full’)) [40]. After GC adjustment, values were transformed to transcripts per million (TPM), to account for the sequencing depth of each sample [42]. Pearson correlations were calculated between normalized read counts of each bin and the following clinical metrics: activated partial thromboplastin time, alanine aminotransferase, albumin, alkaline phosphatase, aspartate aminotransferase, bilirubin, C-reactive protein, creatinine, erythrocytes, hemoglobin, leukocytes, procalcitonin, Quick’s value, thrombocytes, and urea. Correlation values were computed for each patient individually and all samples combined. For the latter , p-values were adjusted for multiple testing using the false discovery rate (FDR) [43]. For metrics with over 50 bins having an FDR < 0.05, the top 50 bins were selected based on the sum of the combined and patient-wise correlation values. The correlates from the identification cohort were evaluated for robustness with the test cohort. This evaluation was based on the correlation value in the test cohort and the mean absolute error (MAE) to a linear model fitted on the identification cohort. Outliers, defined as values outside the identification cohort’s clinical metric data range, were excluded from thew test cohort. Correlates with the strongest correlation and smallest MAE were manually manual inspected to ensure that the correlation was not overestimated by a subset of data points. All correlation analyses were executed with python.

### Statistics

Summary statistics for sepsis infection type and sepsis outcome cohorts were calculated in R (tidytlg; v0.1.4). For numerical metrics, mean with standard deviation, median, range, and interquartile range are reported. Categorical metrics are reported as percentages. Numerical metrics were tested for difference of medians using a two-sided Wilcoxon rank-sum test. Categorical metrics were tested for differences in proportions using a two-sided Fisher’s exact test. The discriminatory power of numerical metrics for the patient groups in the two cohorts was evaluated using the area under the receiver operating characteristic (AUROC) curve calculated in R (pROC; v1.18.0). 95 % confidence interval values of AUROC values are reported and were calculated using DeLong’s method. Differences between ROC curves were tested for significance with the bootstrap method of pROC. Optimal classification thresholds are provided that have been calculated using the Youden criterion.

Optimal classification thresholds derived from ROC analyses were used to categorize patients in the outcome cohort into high-risk or low-risk groups for mortality. Subsequently, time-to-event analyses were conducted to evaluate potential differences between these groups. Cox proportional hazards models were fitted using R, and significance of differences was assessed with pairwise_survdiff function from survminer (v0.4.9) and survival (v3.5.7). Hazard ratios were calculated and tested for statistical significance using the Wald test.

## Results

### Characterization of footprint DNA

Released DNA in the bloodstream gets rapidly degraded by DNA digesting enzymes, like DNase I, generating circulating cell-free DNA (cfDNA). Histones and other DNA binding proteins (DBP), like transcription factors, protect cfDNA from degradation (**Figure 1a**). Footprint DNA fragments originating from DBPs are generally shorter compared to cfDNA from histones, which facilitates specific enrichment and high-throughput sequencing of these fragments. Sequencing reads of footprint DNA are enriched at regulatory genomic sites like promoters, enhancers, transcription factor binding sites (TFBS), at CpG islands or in proximity to transcription start sites (TSS) [12,44] (**Figure 1a, b**, **Supplement Figure 1** and **Supplement Figure 2**). An exemplary genomic region with enrichment of footprint DNA reads is shown for the NOCT gene (**Figure 1b**). Enrichment and sequencing of footprint DNA provides genome-wide information on regulatory interactions between DBPs and the genome, which correlates with the host response of sepsis patients and thus can be used to identify valuable biomarkers. Accordingly, genomic regions with differentially enriched footprint DNA reads, potentially revealing bona fide biomarkers, were inferred from peak calls, while correlations between footprint DNA signals and clinical metrics were identified from genomic partitions (bins) of 100 bp in size (**Figure 1b** zoom).

**Figure 1:**
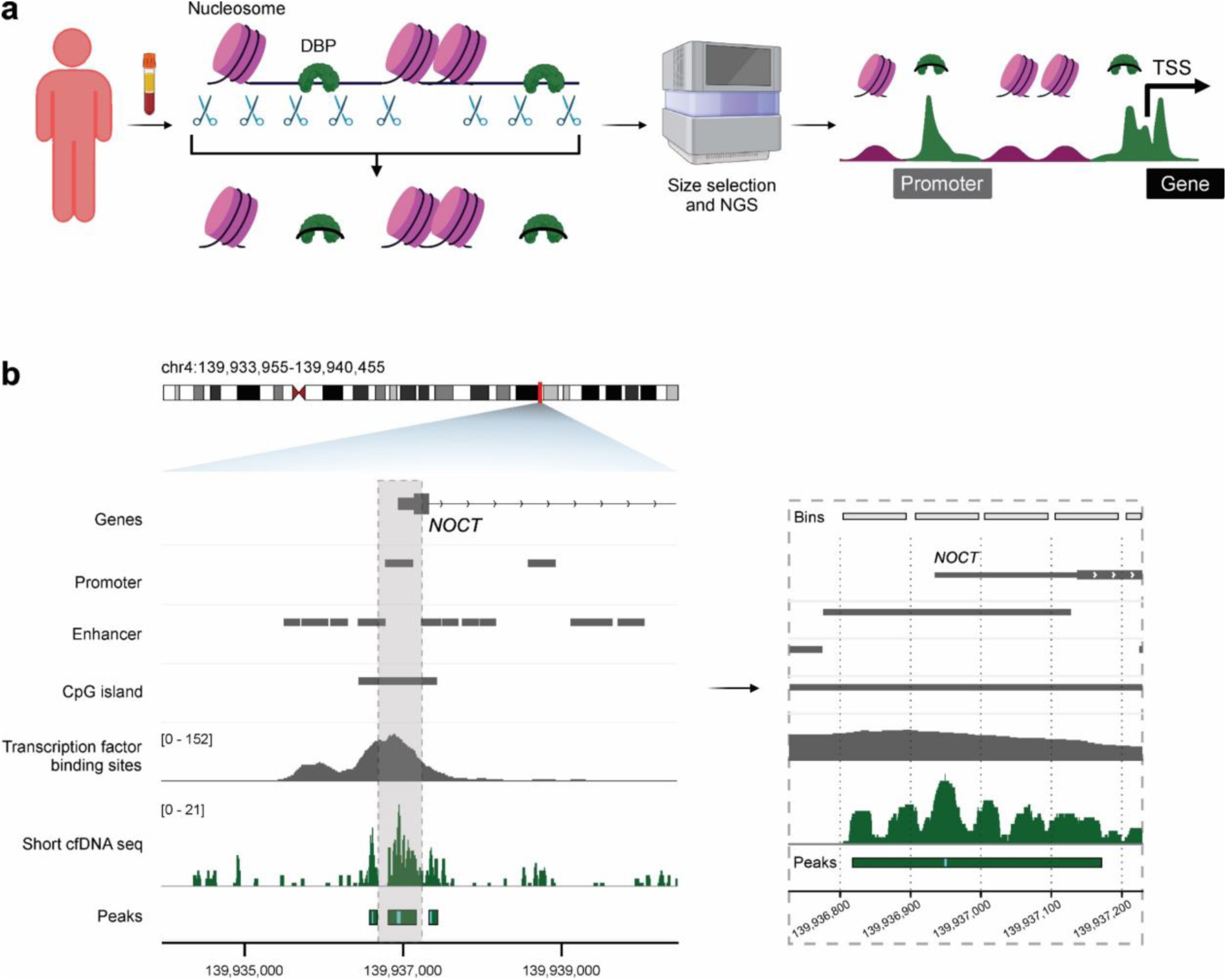
Origin of footprint DNA and genome-wide signal extraction approaches. a) CfDNA is protected from enzymatic digestion in the bloodstream either by histones (purple) or other DNA-binding proteins (DBP, footprint DNA, green). Isolating cfDNA from the plasma of patients, enriching DBP-bound footprint DNA according to size and sequencing it yields amplified signals at regulatory elements such as promoters or transcription start sites (TSS) of genes. Created in BioRender [Sonntag M., 2025, https://BioRender.com/i06x610]. Exemplary footprint DNA read enrichment at the *NOCT* gene promoter, enhancer, CpG island, and various TFBS. Either by peak calling (left) or by dividing the human genome into bins, footprint DNA signals can be extracted and analyzed (right, zoom in).

### Footprint DNA contains information about sepsis patient physiology

Footprint DNA provides genome-wide information about the interaction of regulatory proteins with the genome. Next, we investigate whether this information can be linked to known physiological functions measured by established clinical parameters. Blood and plasma samples were collected from sepsis patients over a period of 21 days for routine clinical measurements and plasma extraction (**Figure 2a**). Patient characteristics and physiological parameter are depicted in **Supplement Table 1** and **Supplement Table 2**. Plasma was used for footprint DNA enrichment, sequencing, and extracted signals from the bin approach were evaluated for correlations to routine laboratory parameters. Parameters were evaluated for liver function (alanine aminotransferase (ALT), albumin, alkaline phosphatase (ALP), aspartate aminotransferase (AST), and bilirubin), kidney function (creatinine and urea), immune response (C-reactive protein (CRP), leukocytes, and procalcitonin (PCT)), blood coagulation system (activated partial thromboplastin time (aPTT), Quick’s value and thrombocytes), as well as oxygen transport (erythrocytes, hemoglobin). The correlations between footprint DNA signals and described metrics were tested both patient-centered and between patients (‘identification’ cohort) as well as validated in completely independent samples (‘test’ cohort).

**Figure 2:**
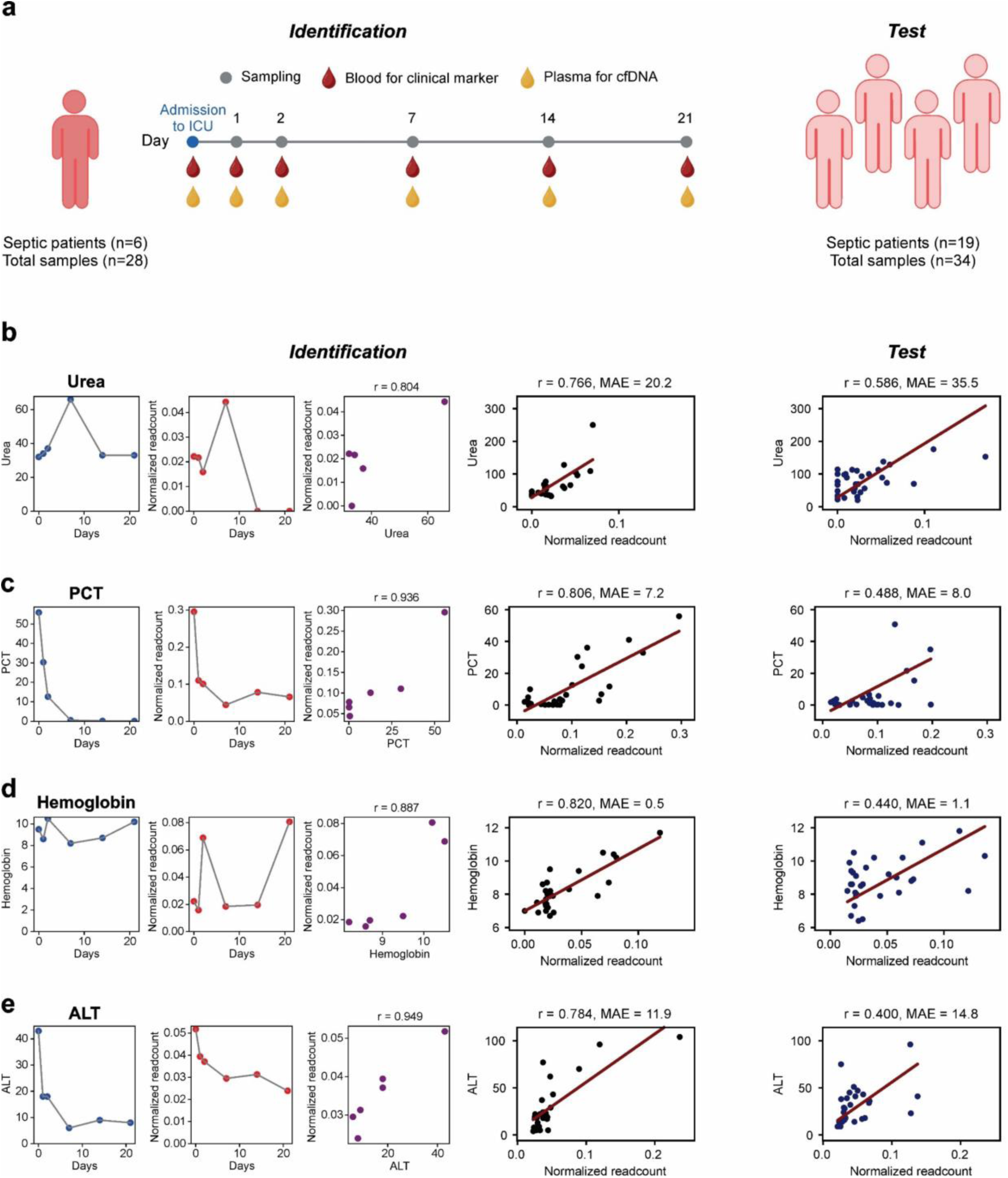
Correlation of footprint DNA signals to physiological markers. a) Experimental design for the identification and verification of footprint DNA correlates. Routine clinical metrics as well as short cfDNA sequencing data were acquired from six septic patients over 21 days (Identification, n=28 samples, 6 individuals). Independent samples from other septic patients were used for validation (Test, n=34 samples, 19 individuals). Created in BioRender [Sonntag M., 2025, https://BioRender.com/w86w257]. b-e) In the identification cohort, footprint DNA markers from the six patients were correlated patient wise to routine clinical metrics (left three plots) and across all patients (second right plot). In the test cohort, independent samples from sepsis patients were used for validation of identified correlation results. Reported correlations are pearson correlation values (r). Adjusted p values of correlations across patients: urea: r=0.766, p=0.0011; PCT: r=0.806, p=0.0163; hemoglobin: r=0.820, p=0.0424; ALT: r=0.784, p=0.0165. The red lines show the fitted linear function based on the identification cohort from which the MAE is calculated. PCT, procalcitonin; ALT, alanine transaminase: MAE, mean absolute error.

For four physiological parameters, significant footprint DNA correlations were identified, including kidney function (urea; **Figure 2b**), immune response (PCT; **Figure 2c**), oxygen transport (hemoglobin; **Figure 2d**), and liver function (ALT; **Figure 2e**). Additionally, weaker correlates for additional liver functions (ALP, AST, and bilirubin; **Supplement Figure 3a-c**) and oxygen transport (erythrocytes; **Supplement Figure 3d**) were also identified. Patient physiology can change dynamically within a few days, leading to an exponential decrease of PCT in patient S33, for example (**Figure 2c**). Within the identification cohort for all six patients combined, footprint DNA signals correlate significantly and strongly with clinical parameters including urea (Pearson r: 0.766) or PCT (Pearson r: 0.806) (**Figure 2b-e**). At the individual patient level, correlations also showed similar trends (**Supplement Figure 4, Supplement Figure 7**). These correlations were then further validated using samples from an independent test cohort (**Supplement Table 3**). Although extent of correlations decreased (Pearson r: 0.586 for urea and 0.440 for hemoglobin) and the MAEs of the linear fits increased, a moderate correlation with the clinical parameters was still observed (**Figure 2b-e** right plot). These results suggest that sequencing footprint DNA can provide semi-quantitative information to describe dynamic physiological changes of clinical metrices, which are used to assess the current status of the patient, e.g., organ function.

### Footprint DNA accurately predicts type of infection in sepsis

In clinical sepsis diagnostic, PCT is routinely measured to monitor bacterial sepsis, with potential use in detecting bacterial sepsis. However, PCT not always provides reliable results to differentiate between bacterial and non-bacterial sepsis. Therefore, our cohort was used to determine whether footprint DNA markers are able to differentiate between sepsis infection types and uninfected controls already at the day of admission to the ICU (**Figure 3a** and **Supplement Table 4**). Bacterial and viral sepsis groups of the cohort do not differ significantly in any of the measured standard clinical parameters (**Supplement Table 5**). In clinical routine, procalcitonin (PCT) is often used as an indicator for bacterial sepsis, however, we found PCT values only significantly different between non-infected postoperative controls (POP) and infected patients, but not between bacterial and viral septic patients (**Supplement Figure 9**). Footprint DNA markers were identified by differential read coverage analysis from the consensus peaks of each condition (**Supplement Figure 10**). The top five differentially enriched regions (DER) per condition and per comparison (POP vs. viral, POP vs. bacterial, bacterial vs. viral; in total 30 DERs) separate the three infection types by the first and second principal components applying a principal component analysis (PCA) (**Figure 3b** and **Supplement Figure 11**). Separation of these groups based on the top 30 DERs from the three comparisons is also accomplished by hierarchical clustering (**Supplement Figure 12** and **Supplement Figure 13a-c**). Differential enrichment analysis for bacterial vs. viral sepsis revealed 220 DER in bacterial and 68 DER in viral sepsis, respectively (**Figure 3c**). Among significantly enriched bacterial DERs, a distinct differential DNA footprint in the promoter region of the *AP1B1* gene could be detected (**Supplement Figure 12**). Among significant viral DERs, differential footprint DNA signals could be exemplarily detected in the promoter of the RNA Polymerase III subunit B (*POLR3B*) or the solute carrier family 30 member 10 (*SLC30A10*), respectively (**Supplement Figure 12**). Receiver operating characteristics (ROC) analysis was used to compare the discriminatory and predictive power of the top DER with PCT. Results show that footprint DNA is a significantly better predictor of sepsis infection type than PCT (area under the ROC curve (AUROC): footprint DNA=1.000 vs. PCT=0.750; p=0.0354) (**Figure 3d**). Additionally, differential enrichment of transcription factor binding motifs analysis was performed to identify regulatory proteins that may be involved in the discrimination of the conditions. In total, 37 significantly enriched transcription factor motifs for bacterial and 186 for viral sepsis were detected, respectively (**Figure 3e,f**). For viral sepsis the most significantly enriched binding motif belongs to SMAD family member 3 (SMAD3), while two Kruppel-like factors (KLF12 and KLF9) were detected among the top 10 most significant motifs in bacterial sepsis (**Figure 3e,f**).

**Figure 3:**
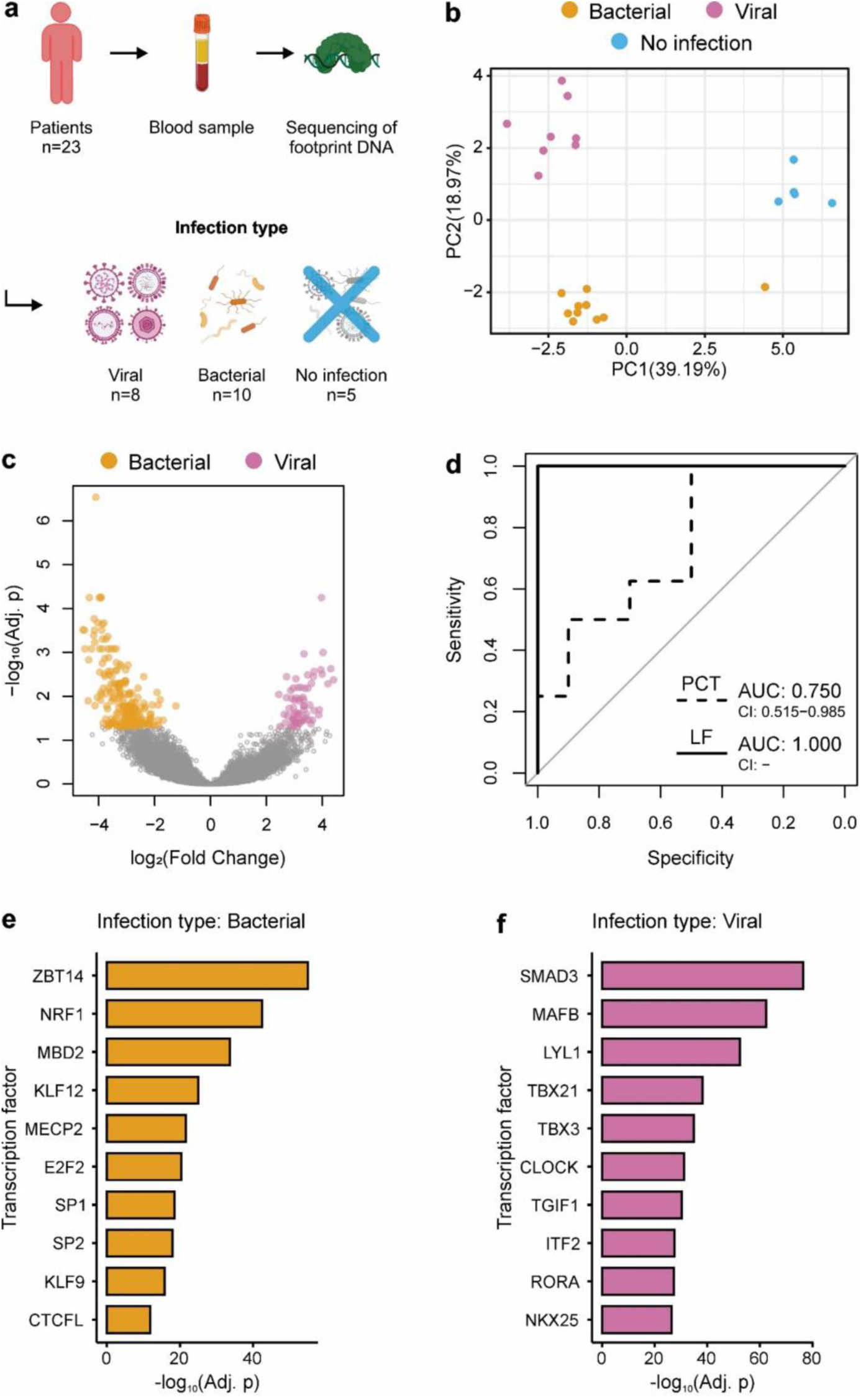
Discrimination of sepsis infection types by footprint DNA. a) Experimental design: For 23 samples (8 viral sepsis patients, 10 bacterial sepsis, 5 non-infected postoperative controls) plasma was prepared, footprint DNA enriched and sequenced. Created in BioRender [Sonntag M., 2025, https://BioRender.com/d50d259]. b) Principal component analysis based on the top 30 identified differentially enriched regions separates viral from bacterial sepsis and from non-infected postoperative controls. c) Differential enrichment analysis volcano plot for the comparison of viral and bacterial sepsis. Differentially enriched regions are colored according to their respective condition (adjusted p-value ≤ 0.05 and |log2 (fold change)| ≥ 0.5). d) Receiver operating characteristics analysis based on the top 10 identified Liquid Footprinting (LF) biomarkers from c) and for the routine clinical parameter PCT (95% confidence intervals are included for AUROC values; p=0.0354). e) and f) Top 10 most significant differentially enriched transcription factor motifs in bacterial sepsis (e) and in viral sepsis (f).

### Footprint DNA accurately predicts early death in sepsis

Prognostic biomarkers for the prediction of outcome in sepsis can be crucial to administer the most suitable treatment options and to reduce adverse outcomes. We therefore defined a cohort of severe sepsis cases (‘outcome’ cohort) including 22 sepsis survivors (survival > 28 days) and 21 early death sepsis cases (death < 3 days). Based on the outcome cohort, we aim to identify footprint DNA biomarkers for identification of early death in sepsis (**Figure 4a** and **Supplement Table 7**). The outcome cohort groups differ significantly in eight of the 27 measured clinical metrics, with the SOFA score showing the lowest p value and the highest AUROC of 0.76 (**Supplement Table 8**). Differential enrichment analysis identified one DER for early death and seven for sepsis recovery, respectively (**Figure 4b**). Based on the top 5 footprint DNA signals per condition, early death and recovery from sepsis could be separated by the second principal component from PCA (**Figure 4c**), as well as by hierarchical clustering (**Supplement Figure 13d**). ROC analysis of the top 10 footprint DNA signals and the most established clinical metric SOFA yielded a significantly better predictive power for the footprint DNA biomarkers (AUROC: footprint DNA=0.983 vs. SOFA=0.760; p=0.002; **Figure 4d**). Additionally, time-to-event analysis revealed that the low-risk group identified by footprint DNA biomarkers had a significantly lower hazard ratio (HR) for death compared to the group identified by the SOFA score (HR: footprint DNA=0.042 (95 % confidence interval: 0.009-0.186), SOFA=0.364 (95 % confidence interval: 0.148-0.896); p=0.015; **Supplement Figure 14**). Differential enrichment analysis of transcription factor binding motifs identified 123 significantly enriched transcription factor motifs for early death and two for sepsis recovery, respectively (**Figure 4e,f**). For recovery in sepsis, the top binding motif reflects the transcription factor BTB domain and CNC homolog 1 (BACH1) (**Figure 4e,f**).

**Figure 4:**
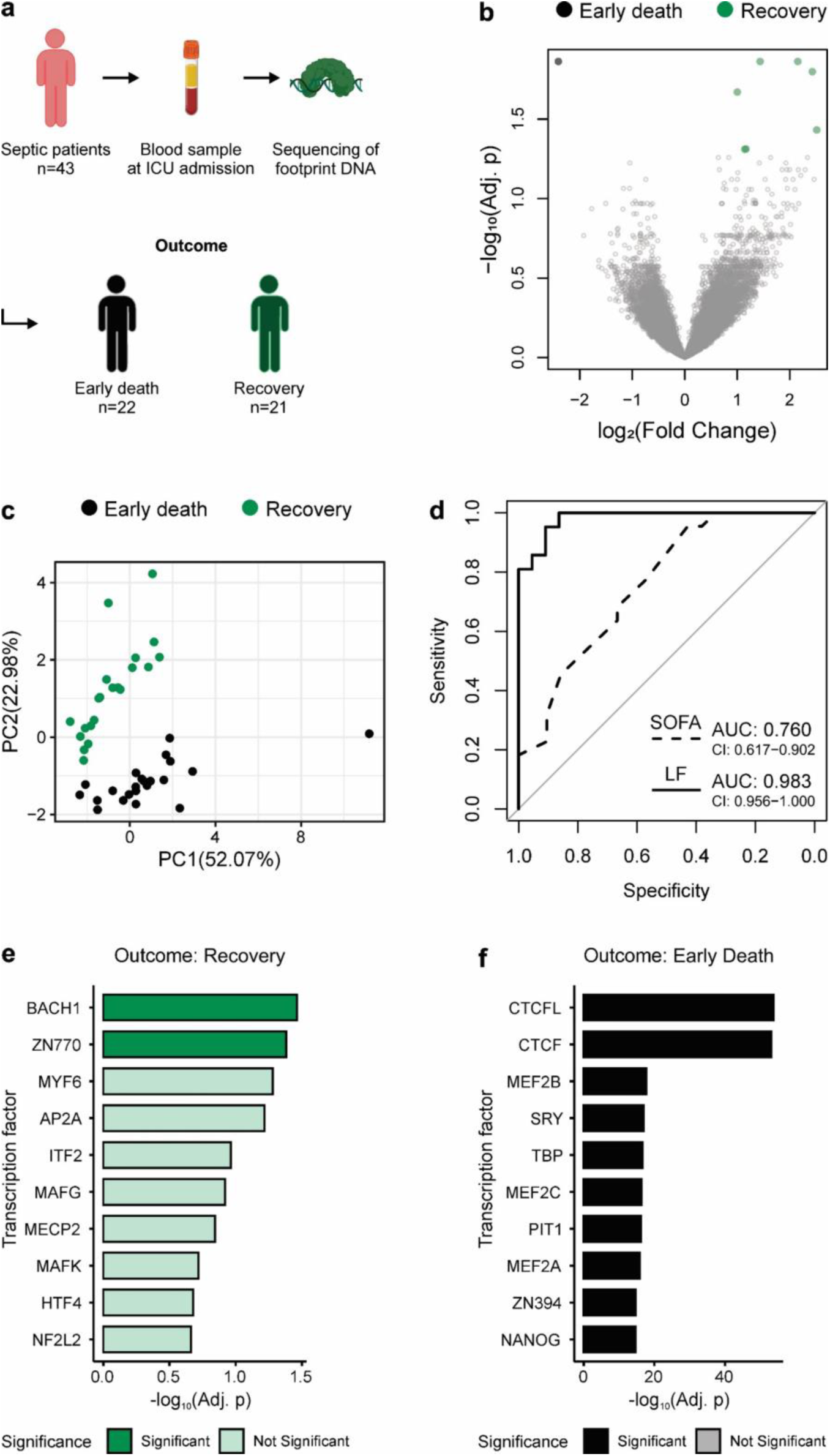
Discrimination of sepsis outcome by footprint DNA. a) Experimental design: From 43 septic patients samples (22 early death sepsis patients and 21 recovery patients) plasma was obtained, footprint DNA enriched and sequenced. Created in BioRender [Sonntag M., 2025, https://BioRender.com/b36q835]. b) Differential enrichment analysis volcano plot for the comparison of early death and recovery in sepsis. Differentially enriched regions are colored according to their respective condition (adjusted p-value ≤ 0.05 and |log2 (fold change)| ≥ 0.5). c) Principal component analysis based on the top 10 identified differentially enriched regions separates early death from recovery in sepsis. d) Receiver operating characteristics analysis based on the top 10 identified Liquid Footprinting (LF) biomarkers from b) and for the routine clinical metric SOFA (95% confidence intervals are included for AUROC values; p=0.002). e) and f) Top 10 most significant differentially enriched transcription factor motifs in recovery (e) and early death in sepsis (f).

## Discussion

Sepsis remains a global health threat, in part due to the lack of specific diagnostic tools to assess the condition of septic patients. Monitoring the host immune response in sepsis offers valuable insights but is currently underutilized in clinical practice [45]. Several approaches aim to utilize host immune response information, for example gene expression data for pathogen detection or miRNA expression data for host response characterization [46,47]. However, dedicated sepsis biomarkers could facilitate early diagnosis, guide anti-infective therapy and track the severity of sepsis over time [48]. Accordingly, our approach aims to identify dedicated sepsis biomarkers based on cell-free DNA that capture the host response on the basis of a non-invasive liquid biopsy.

Footprint DNA sequencing provides genome-wide information on the interaction between regulatory proteins and the genome, hence signals mainly occur at regulatory genomic elements such as promoters or TFBS (**Figure 1** and **Supplement Figure 1**, **Supplement Figure 2**). Individual footprint signals correlate to clinical, physiological parameters, such as liver (for ALT, r = 0.784) and kidney (for urea, r = 0.766 (**Figure 2**). These footprint DNA markers capture rapid changes of the clinical metrics on single patient level as well as across all patients (**Supplement Figure 4**, **Supplement Figure 7**, **Supplement Figure 8**), underlining that the dynamic in patient’s physiology can be assessed during disease progression, i.e., during a stay on the ICU. Despite the moderate correlation strength observed in the test cohort, the identified correlates suggest that DNA footprinting provides semi-quantitative data about dynamic changes in the physiological state of a patient. Footprint DNA allows body- and genome-wide the monitoring of different organ systems beyond the hematologic system by capturing the dynamics of clinical metrices. These results also imply that a DNA-based liquid biopsy marker can deliver physiological insights about the patient’s status, similar to gene expression data from sources such as whole blood [49]. While this gene expression analysis only depicts signal mainly from the hematopoietic system, footprint DNA is capable deliver additionally information system-wide, e.g., from different organ systems or tumors. Advanced models that integrate multiple footprint DNA signals might further increase correlation and therefore the prediction of established clinical metrics. However, directly replicating well-established, reliable, and inexpensive clinical metrics offers limited added value. Furthermore, the development of such models is limited by the comparably small cohort size.

Inadequate treatment is associated with increased mortality in septic patients, while adequate and early administration of anti-infective treatment leads to improved survival chances [50,51]. Based on the top 30 DERs, we could distinguish non-infected controls from bacterial and viral sepsis (**Figure 3b**). and with the top 10 DERs discriminate viral sepsis cases from bacterial cases with an AUROC of 1.000, outperforming all clinical routine biomarkers for infection including PCT (AUROC = 0.750) (**Figure 3c,d**; **Supplement Table 6**). Among significant DERs enriched in bacterial sepsis, a footprint signal at the promoter of AP1B1 was detected. AP1B1 is located at the Golgi complex to mediate recruitment and sorting of intracellular processes. It is a known regulator of Stimulator of interferon genes (STING) and thereby the cyclic GMP–AMP synthase (cGAS)-STING pathway. STING functions as a receptor for bacterial cyclic dinucleotides and small molecules that activates immunity during bacterial infection [52]. An enhancer in the AKT serine/threonine kinase 3 (AKT3) gene body was also identified as DER. AKT3 is known to be manipulated by bacterial infection and takes part in immune cell signaling including macrophages [53,54]. For viral sepsis, a significant DER in the promoter of POLR3B was detected. POLR3B is a part of the RNA polymerase III complex and senses common DNA viruses, such as cytomegalovirus, vaccinia, herpes simplex virus-1 and varicella zoster virus. This polymerase detects and transcribes viral genomic regions to generate AU-rich transcripts that bring to the induction of type I interferon [55,56]. Based on the transcription factor motif analysis, transcription factors enriched in either bacterial or viral sepsis were identified (**Figure 3e,f**). While the transcription factor SMAD3 is known to be activated during viral infection[57], the family of KLF transcription factors are induced during bacterial infection [58,59]. Also, footprint DNA sequencing outperforms common clinical routine metrics that were summarized in a recent review by Ahuja et al. [60]. Exemplarily for PCT, studies are described to reach an AUROC of 0.85 or lower discriminating bacterial sepsis, which is in concordance with our infection cohort for PCT [61]. Other approaches using either miRNA signatures for host response profiling reach AUROC of 0.90 and 0.83 for bacterial or viral vs. non-infection, respectively [62], while integrated host response analysis for microbe detection reaches 0.96 [63]. Current clinical metagenomics detects microbial-derived cfDNA and is therefore limited in their analysis to bacteria and DNA viruses [4,11]. With footprint DNA biomarkers, also RNA-based microorganisms, e.g., RNA viruses, should be detected based on the host response analysis. Overall, our results show that footprint DNA biomarkers have the potential to discriminate sepsis infection types at an early time point solely on the host response, thereby delivering important information for adequate treatment decisions.

In addition, we used ten footprint DNA markers to prognostically differentiate between early death in sepsis and recovery cases on the day of ICU admission (**Figure 4a-c**). With an AUROC of 0.983, footprint DNA marker’s discriminatory power is significantly better than that of clinical disease severity scoring tools such as the SOFA score (AUROC = 0.760) (**Figure 4d**, **Supplement Table 9**). The SOFA score for prediction of early death in critically ill patients had a sensitivity of 80% in a study by Ferreira et al. [64]. Also, other clinical parameters like lactate (AUROC = 0.66), lactate and SOFA (AUROC = 0.679 for initial sampling), presepsin (91.5 % sensitivity 28-day mortality) or a combination of several biomarkers including interleukin 6 and PCT (AUROC = 0.823) did not perform as promising as footprint DNA markers [65–68]. Recent approaches utilizing machine learning models with readily available clinical data were able to reach AUROCs between 0.83 to 0.89 depending on study and used data [69–71]. The top TF motifs enriched in early death cases, include different cell fate determining transcription factors including myocyte enhancer factor 2 (MEF2) and Nanog homeobox (NANOG). However, a direct causality to sepsis severity or survival remains elusive, yet.

Footprint DNA sequencing offers promising results for the quantitative assessment of the dynamic physiological changes and functionality of organ systems and for the prediction of the infection type and outcome of sepsis. At the current stage, the main limitation of this proof-of-concept study is its small sample size of the analyses. Follow up studies are essential to verify the results. The complete procedure for footprint DNA sequencing and quantification has currently a turnaround time of two days, which is valuable time for critically ill patients. Therefore, a future improvement of the approach will involve the quantification of the most promising footprint DNA markers in a targeted approach to shorten the turnaround time for clinical applicability. Nonetheless, footprint DNA sequencing represents a promising new platform to identify liquid biopsy biomarkers, here presented for application in sepsis diagnostics. By combining the evaluation of the host response using footprint DNA with the evaluation of pathogens using clinical metagenomics, a more holistic assessment of sepsis could be achieved in the future. Footprint DNA biomarkers might be valuable not only for sepsis but also in other complex diseases including autoimmune diseases, cancer or other related fields [16].

## Contributors

MS and KS conceptualized the presented ideas with input from JM. MS performed all laboratory experiments. JM performed computational and statistical analysis of all data with help of AvH. Further interpretation and analysis of the results was carried out by JM, MS and KS. JM prepared the figures. TB, SOD as well as the GAIN-CARE collaborators recruited the patients’ samples used for this study. MS, JM, and KS wrote and revised the manuscript. KS supervised the project. All authors reviewed and approved the manuscript.

## Data sharing statement

The raw high-throughput sequencing data used in this study have been submitted to the NCBI BioProject database (https://www.ncbi.nlm.nih.gov/bioproject) under the accession number PRJNA1134400. Custom data analysis code created for this work is available on GitHub (https://github.com/janmueller17/short_cfDNA_for_sepsis).

## Supporting information

Publication licenses

## Data Availability

https://www.ncbi.nlm.nih.gov/bioproject/PRJNA1134400

## Acknowledgements

We want to thank all clinicians, nurses and patients of the participating intensive care units of both clinical studies.

## Supplementary Data

**Supplement Figure 1:**
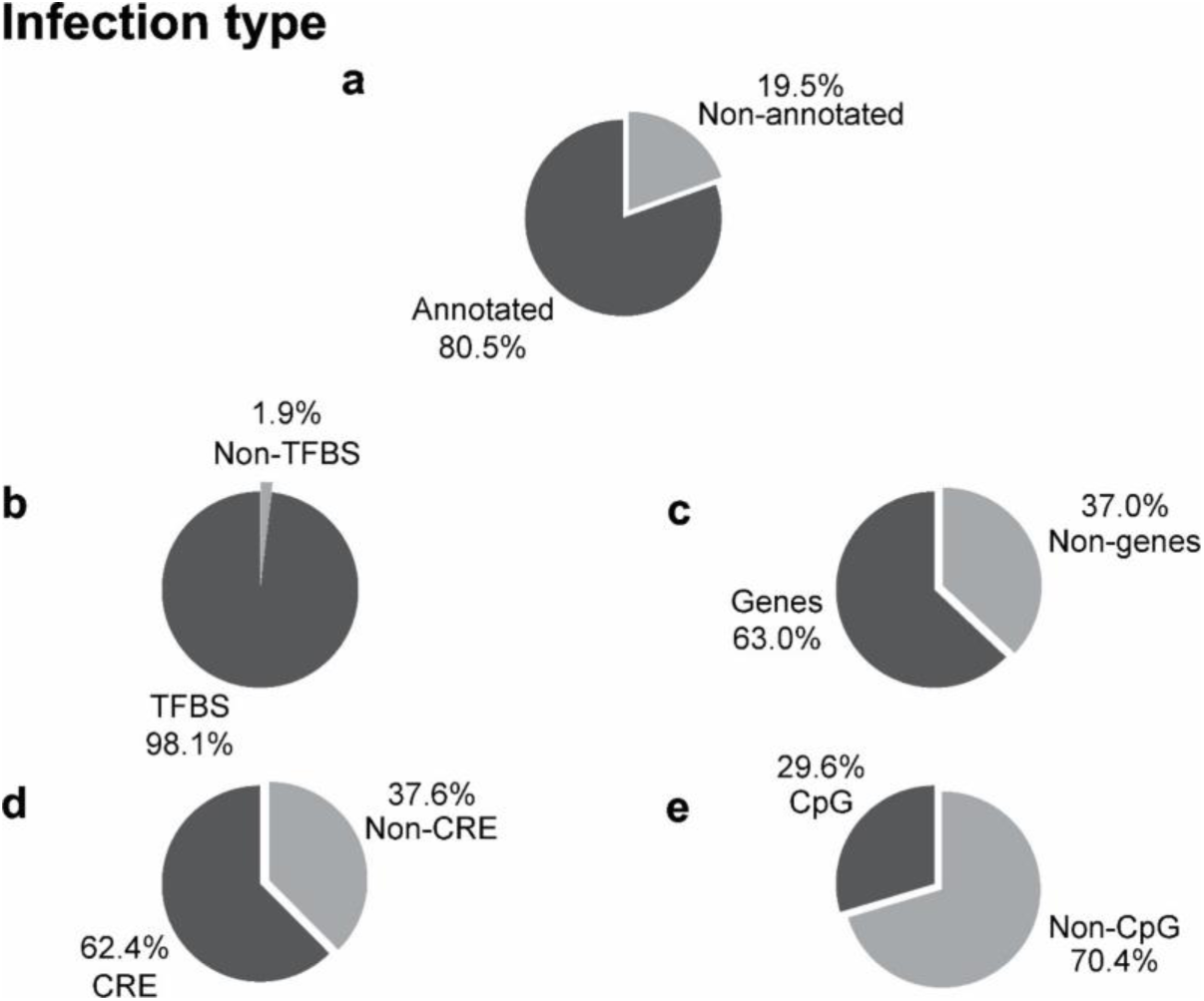
Characterizing of footprint DNA signals in regulatory elements for the infection type cohort. Obtained peaks from footprint DNA signals were mapped against the human genome (a). All annotated peaks were further mapped against annotated regulatory elements including transcription factors bindings sites (TFBS, b), genes (c), cis-regulatory elements (CRE, d) and CpG islands (e).

**Supplement Figure 2:**
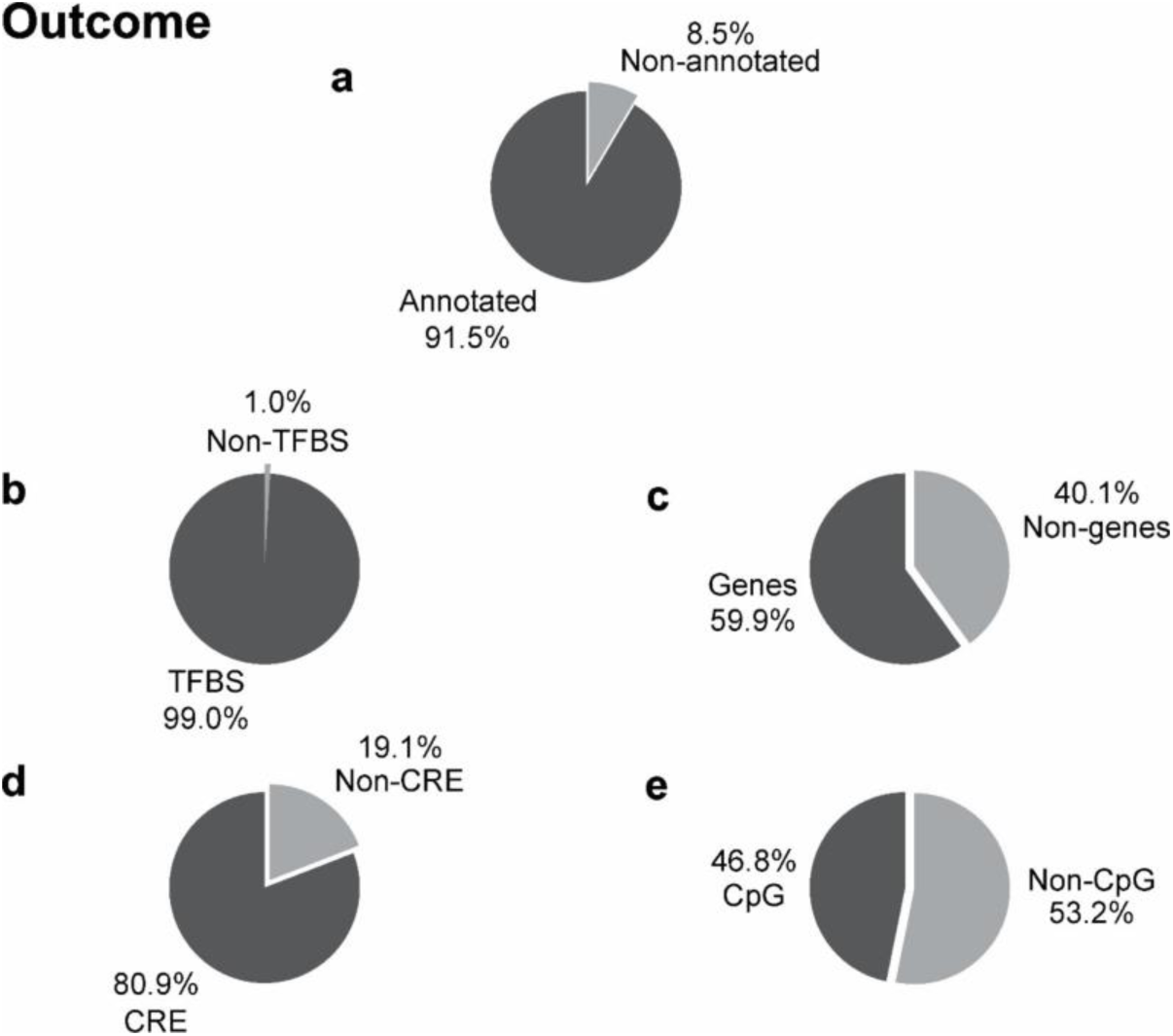
Characterizing of footprint DNA signals in regulatory elements for the outcome cohort. Obtained peaks from footprint DNA signals were mapped against the human genome (a). All annotated peaks were further mapped against annotated regulatory elements including transcription factors bindings sites (TFBS, b), genes (c), cis-regulatory elements (CRE, d) and CpG islands (e).

**Supplement Figure 3:**
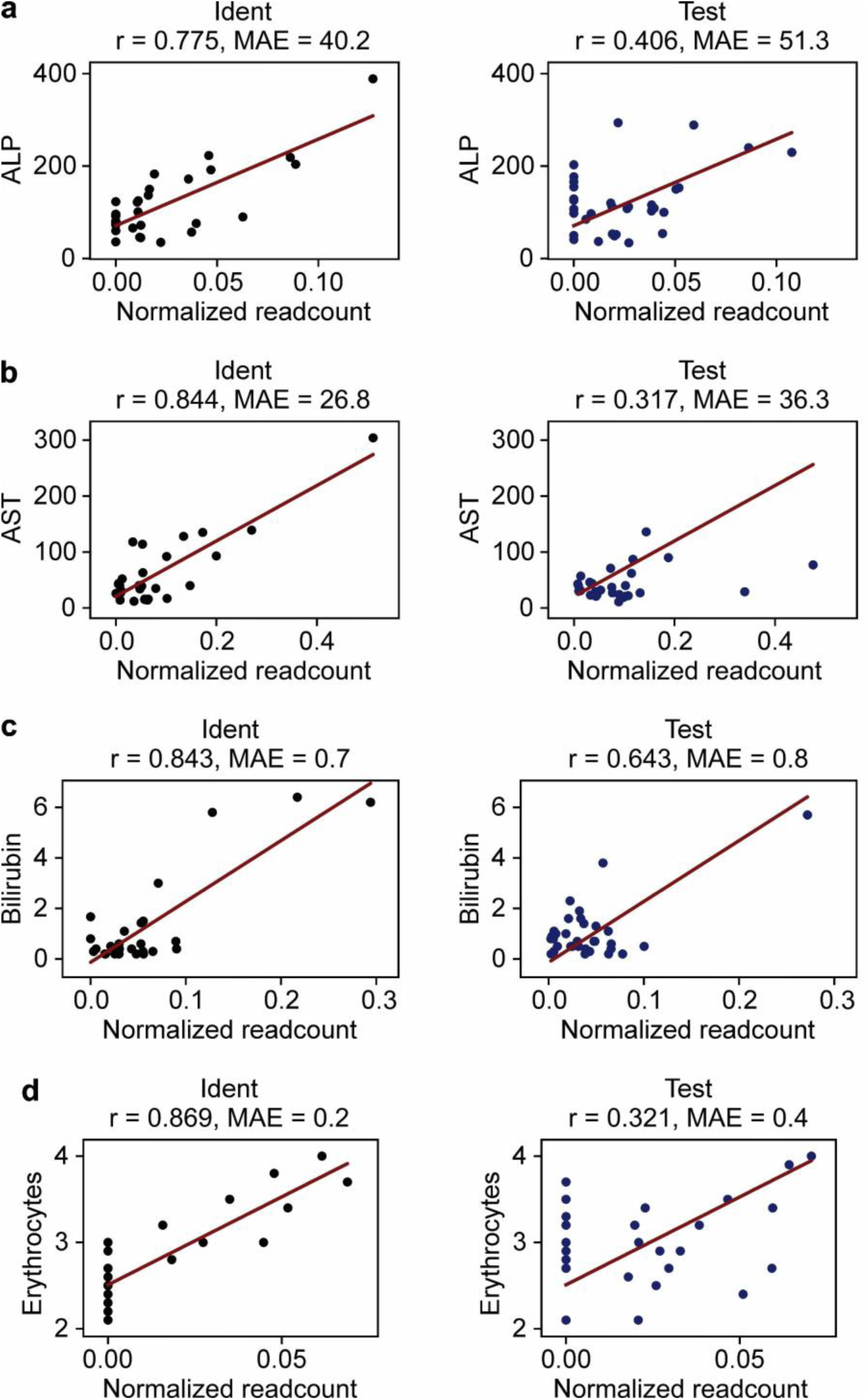
Correlation of footprint DNA signals to additional physiology markers. Routine clinical metrics as well as short cfDNA sequencing data were acquired from six septic patients over a 21 days timespan (Identification, n=28 samples). Independent samples from other septic patients were used for validation (Test, n=34 samples). In the identification cohort, footprint DNA markers from the six patients were correlated to routine clinical metrics across patients (left plot). In the test cohort, independent samples from sepsis patients were used for validation of identified correlation results (middle plot). Correlation of footprint DNA signals to a) Alkanine phosphatase (ALP), b) Aspartate aminotransferase (AST), c) Bilirubin, d) Erythrocytes. Reported correlations are pearson correlation values (r). Adjusted p values of correlations across patients: ALP: r=0.775, p=0.0011; AST: r=0.844, p=0.0163; bilirubin: r=0.843, p=0.0424; Erythrocytes: r=0.869, p=0.0165. The red lines show the fitted linear function based on the identification cohort from which the MAE is calculated. MAE, mean absolute error.

**Supplement Figure 4:**
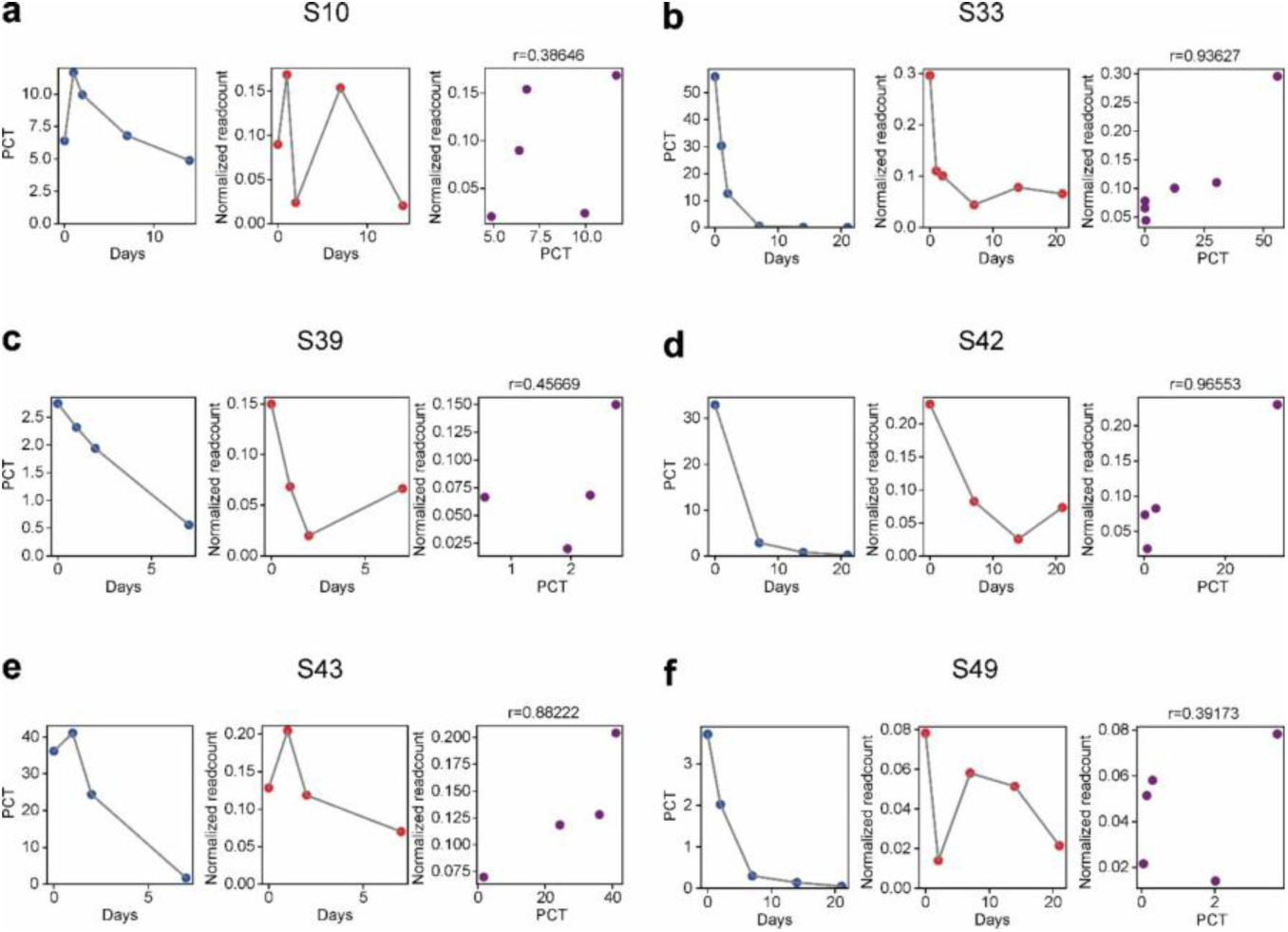
Correlation of footprint DNA markers to the physiological parameter PCT for single patients. a-f) Footprint DNA markers were analyzed for correlation to clinical metric PCT for all patients in the physiological cohort identification set. In the left and middle panel, PCT and normalized readcounts for the footprint, respectively, is depicted over time. The correlation of footprint DNA normalized counts and the clinical metric PCT is shown in the right panel. Results for patients are depicted the following: a) Patient S10, b) Patient S33, c) Patient S39, d) Patient S42, e) Patient S43 and f) Patient S49. PCT, procalcitonin.

**Supplement Figure 5:**
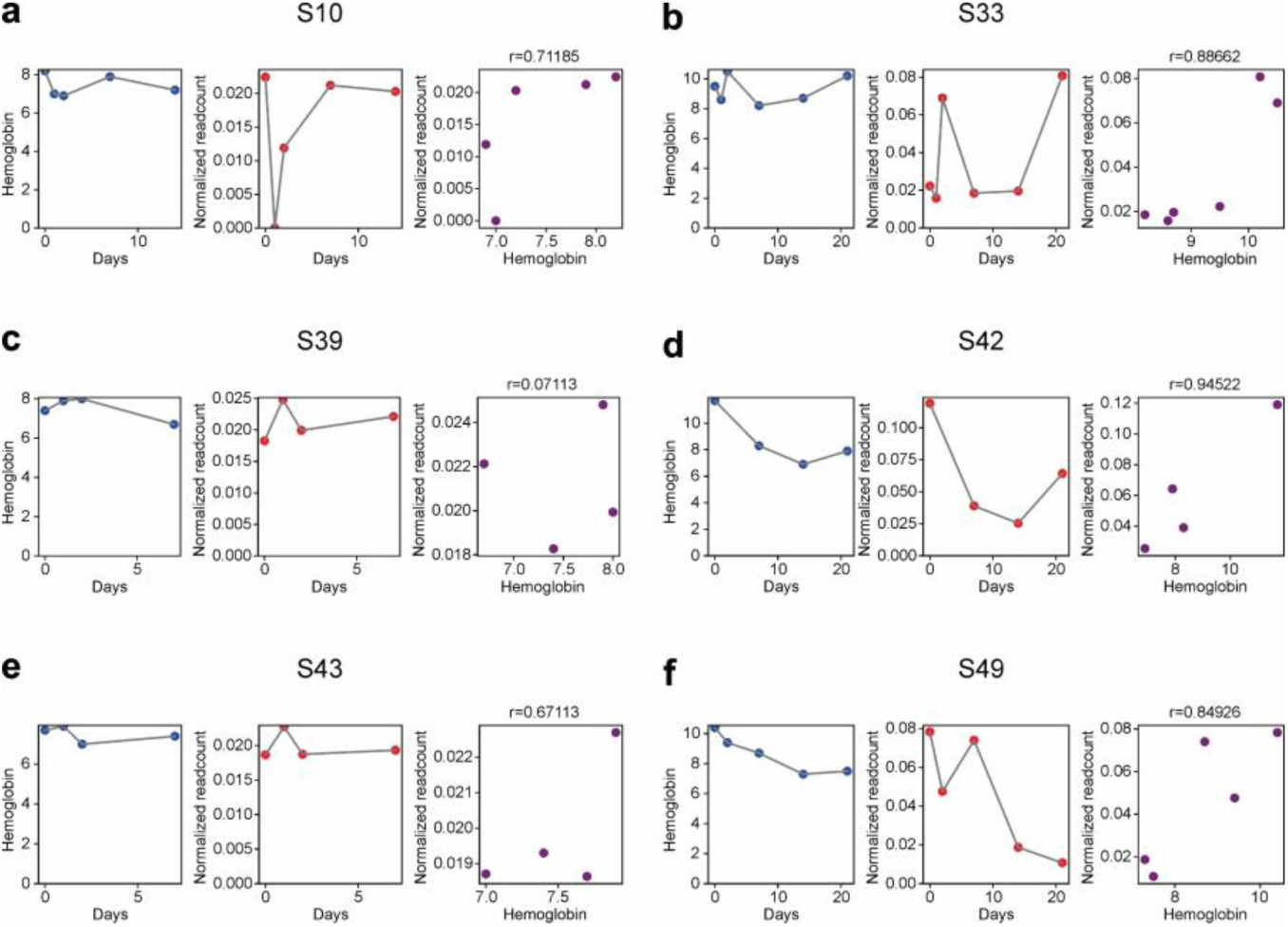
Correlation of footprint DNA marker to the physiological parameter hemoglobin for single patients. a-f) Footprint DNA markers were analyzed for correlation to the clinical metric hemoglobin for all patients in the physiological cohort identification set. In the left and middle panel, hemoglobin and normalized readcounts for the footprint, respectively, is depicted over time. The correlation of footprint DNA normalized counts and the clinical metric hemoglobin is shown in the right panel. Results for patients are depicted the following: a) Patient S10, b) Patient S33, c) Patient S39, d) Patient S42, e) Patient S43 and f) Patient S49.

**Supplement Figure 6:**
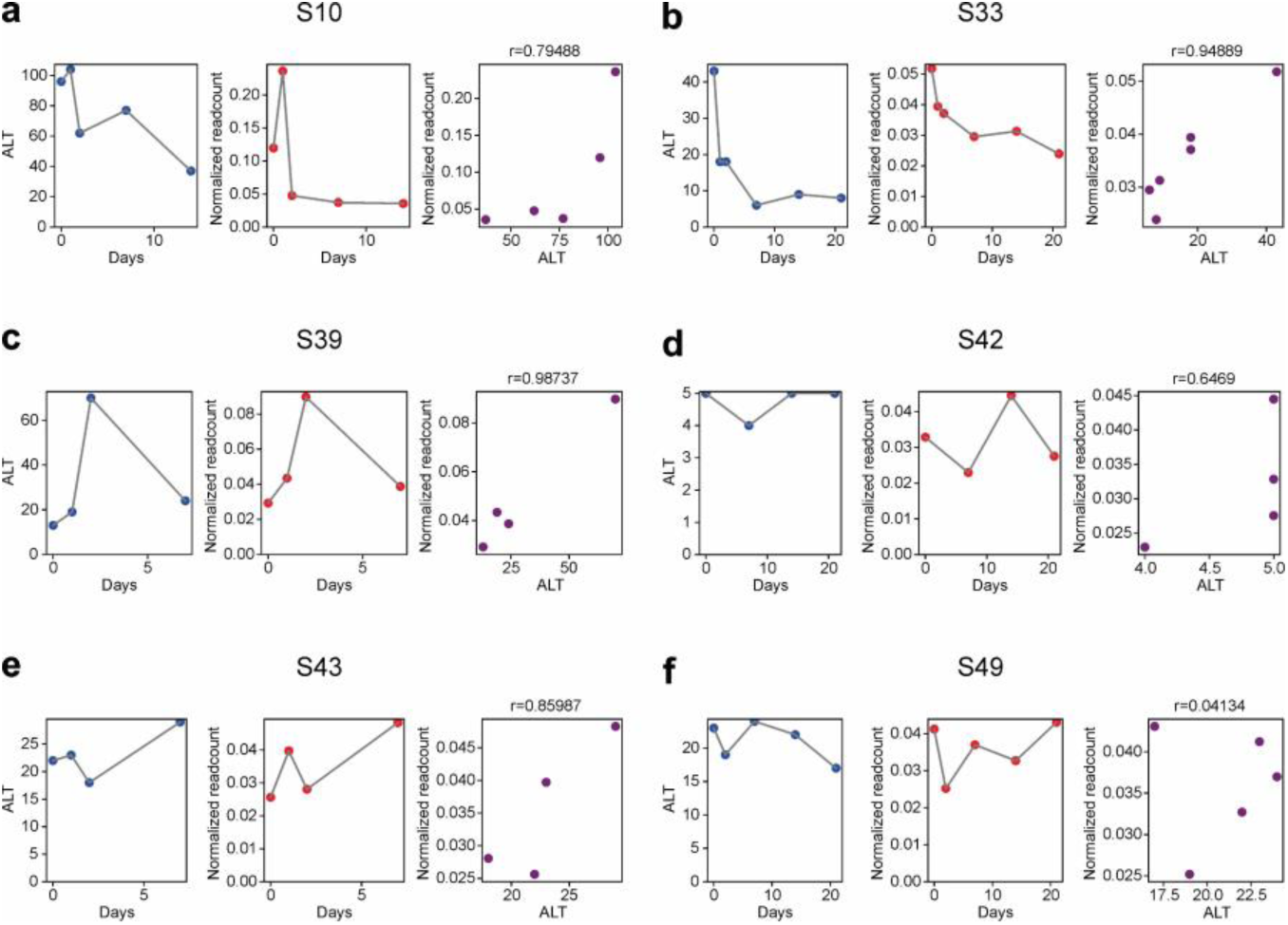
Correlation of footprint DNA marker to the physiological parameter ALT for single patients. a-f) Footprint DNA markers were analyzed for correlation to clinical metric ALT for all patients in the physiological cohort identification set. In the left and middle panel, ALT and normalized readcounts for the footprint, respectively, is depicted over time. The correlation of footprint DNA normalized counts and the clinical metric ALT is shown in the right panel. Results for patients are depicted the following: a) Patient S10, b) Patient S33, c) Patient S39, d) Patient S42, e) Patient S43 and f) Patient S49. ALT, alanine aminotransferase.

**Supplement Figure 7:**
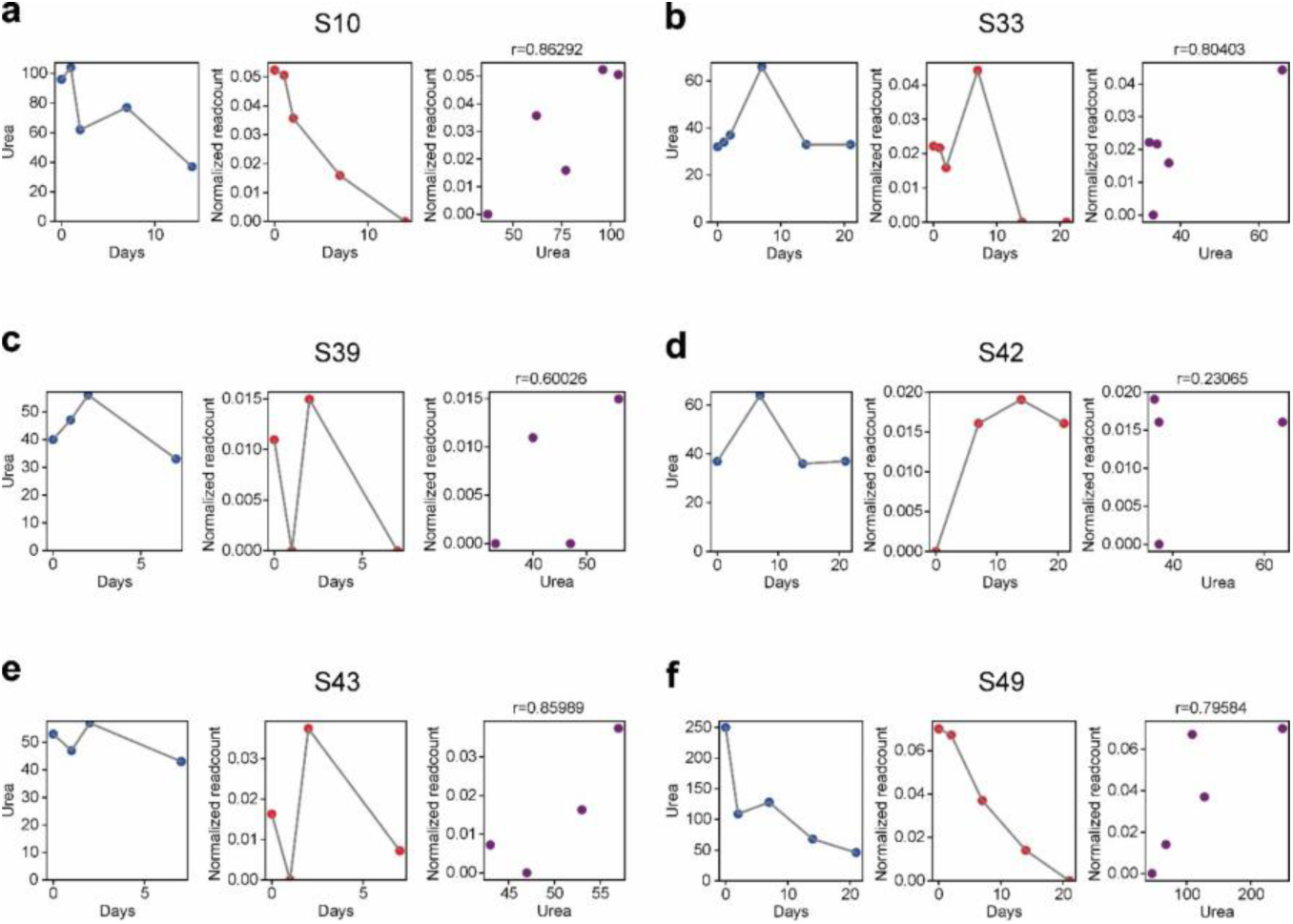
Correlation of footprint DNA marker to the physiological parameter urea for single patients. a-f) Normalized readcounts of footprint DNA markers were analyzed for correlation to the clinical metric urea for all patients in the physiological cohort identification set. In the left and middle panel, urea and normalized readcounts for the footprint, respectively, is depicted over time. The correlation of footprint DNA normalized counts and clinical metric urea is shown in the right panel. Results for patients are depicted the following: a) Patient S10, b) Patient S33, c) Patient S39, d) Patient S42, e) Patient S43 and f) Patient S49.

**Supplement Figure 8:**
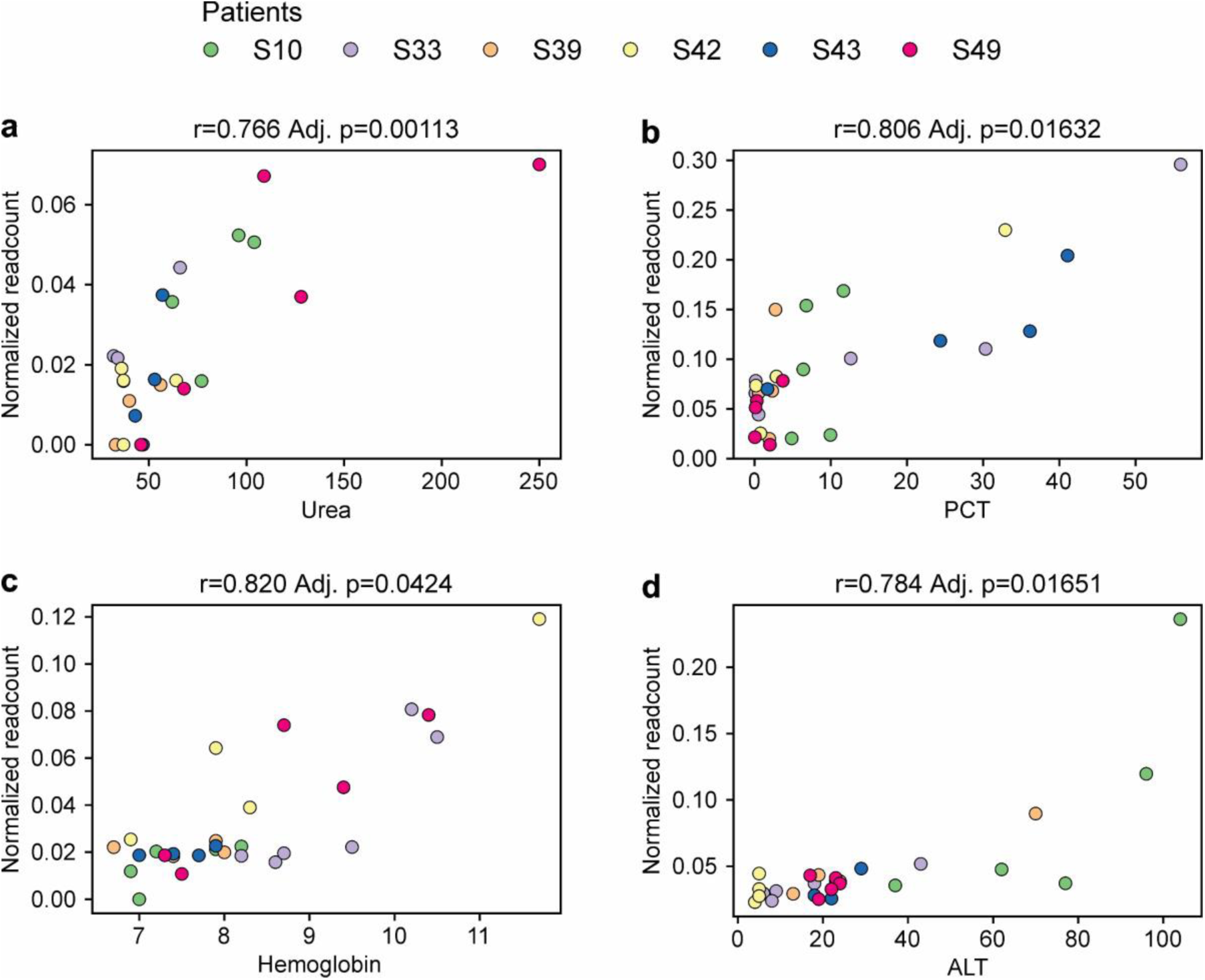
Patient-wide correlation of footprint DNA markers to physiological parameters. a-d) Normalized readcounts of footprint DNA markers were analyzed for correlation to each clinical metric combined for all patients in the physiological cohort identification set. Results for clinical metrics are depicted the following: a) urea, b) PCT, c) hemoglobin and d) ALT. PCT, procalcitonin; ALT, alanine aminotransferase.

**Supplement Figure 9:**
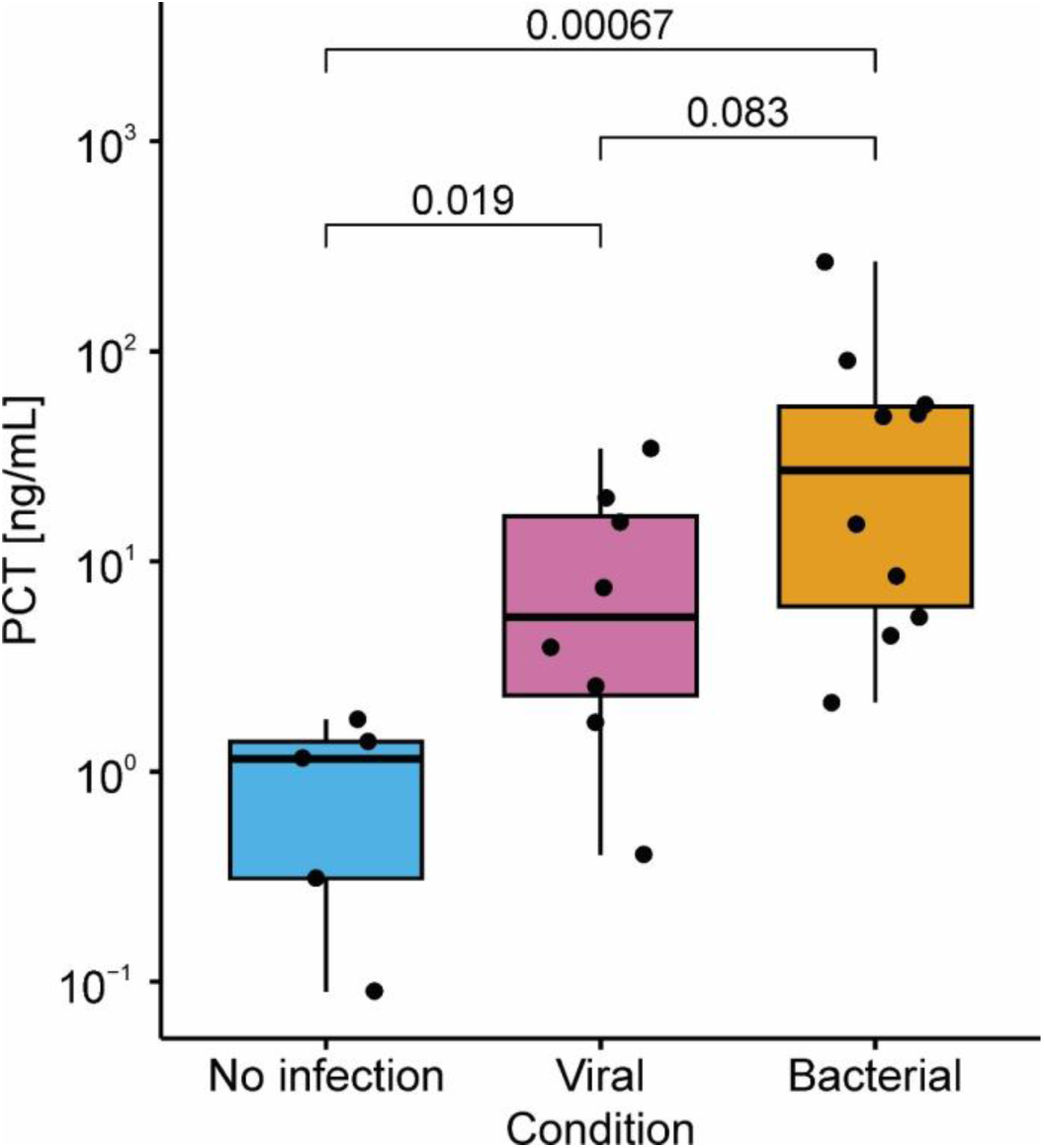
PCT concentration in the infection type cohort. The concentration of Procalcitonin (PCT) was measured for each sample and categorized to non-infection, postoperative controls (blue), viral sepsis (pink) and bacterial sepsis (yellow). Statistical testing was performed using a two-sided Wilcoxon rank-sum test. Resulting p-values are indicated above the brackets for each comparison.

**Supplement Figure 10:**
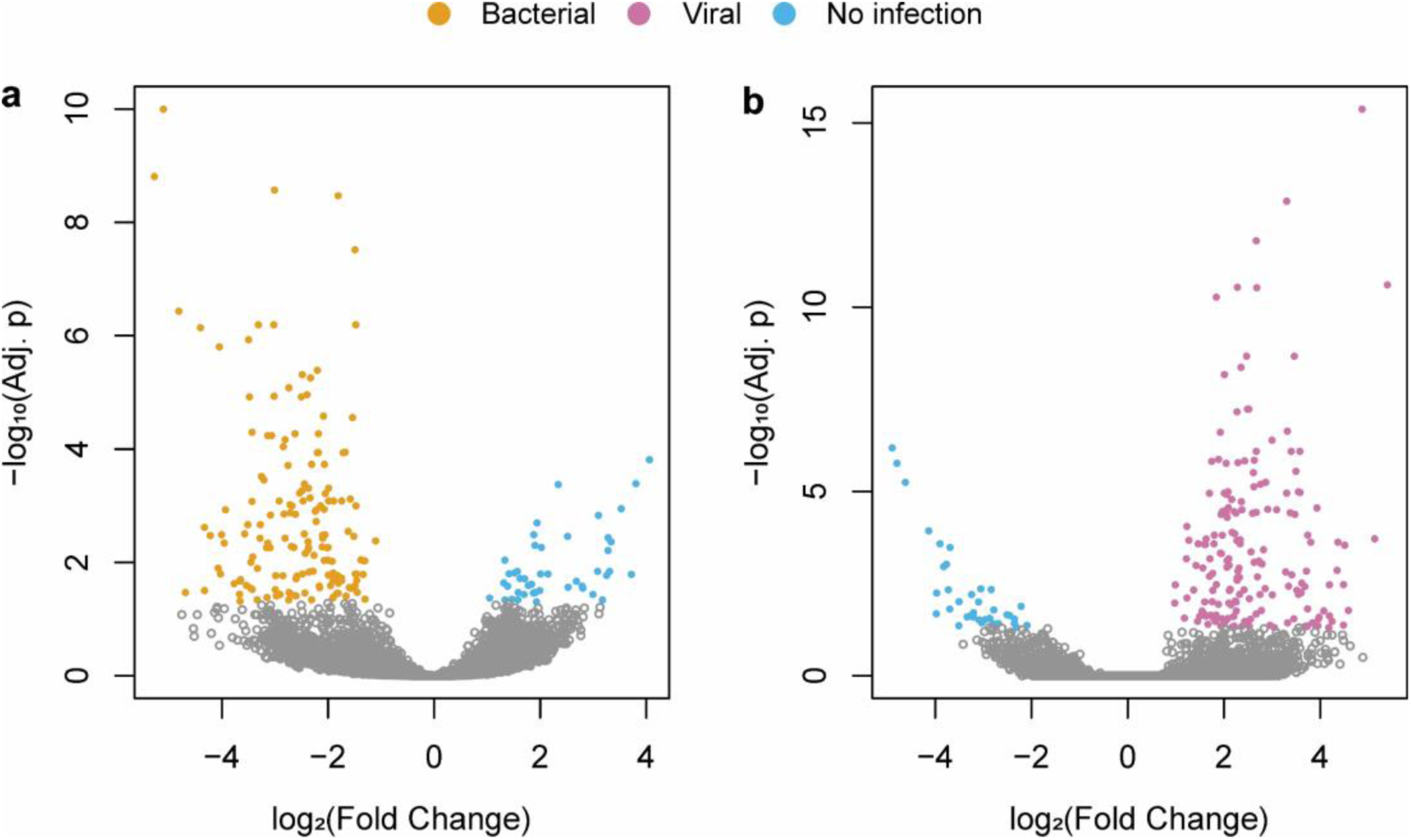
Differential enrichment analysis for non-infection controls and sepsis samples with viral or bacterial sepsis, respectively. Differentially enriched regions (DERs) were analyzed between two conditions for enriched transcription factor binding sites (log10 Adj. p-value ≤ 0.05 and log2 Fold change ≥ 2). Significant results are colored and depicted in a Volcano plot. a) non-infection controls (blue) were analyzed against bacterial sepsis cases (yellow) b) non-infection controls (blue) were analyzed against bacterial sepsis cases (pink).

**Supplement Figure 11:**
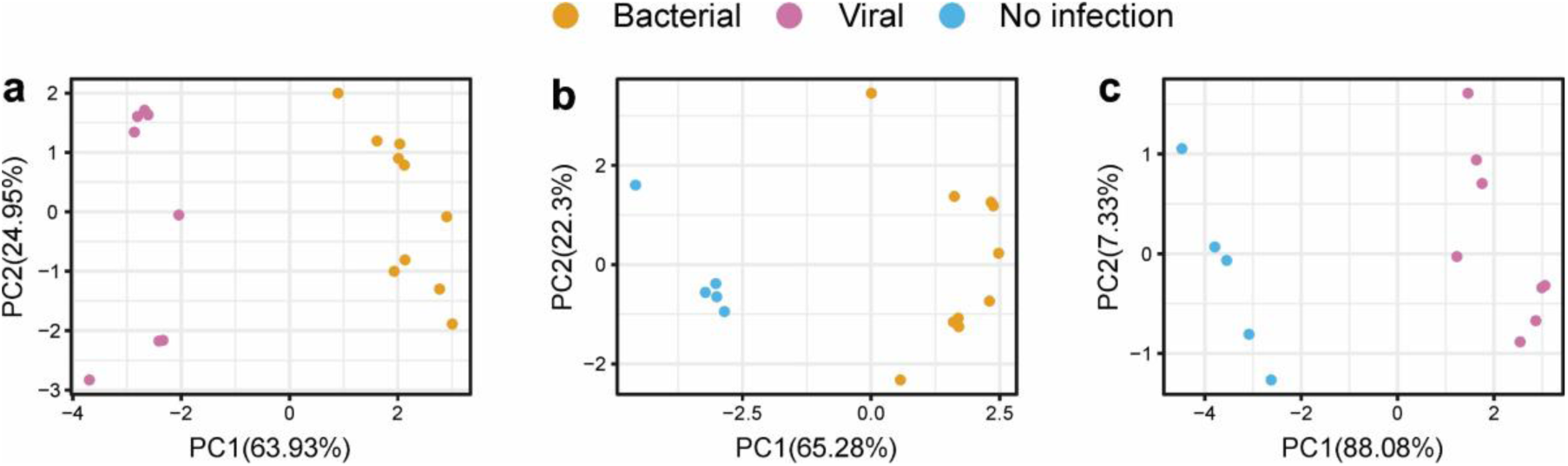
Discrimination of non-infection, postoperative controls, viral and bacterial sepsis via PCA. Based on the top 5 differential enriched regions (DERs) per condition, principal component analysis (PCA) was performed. The following conditions of the infection type cohort were investigated a) bacterial sepsis against viral sepsis b) non-infection control against bacterial sepsis c) non-infection control against viral sepsis.

**Supplement Figure 12:**
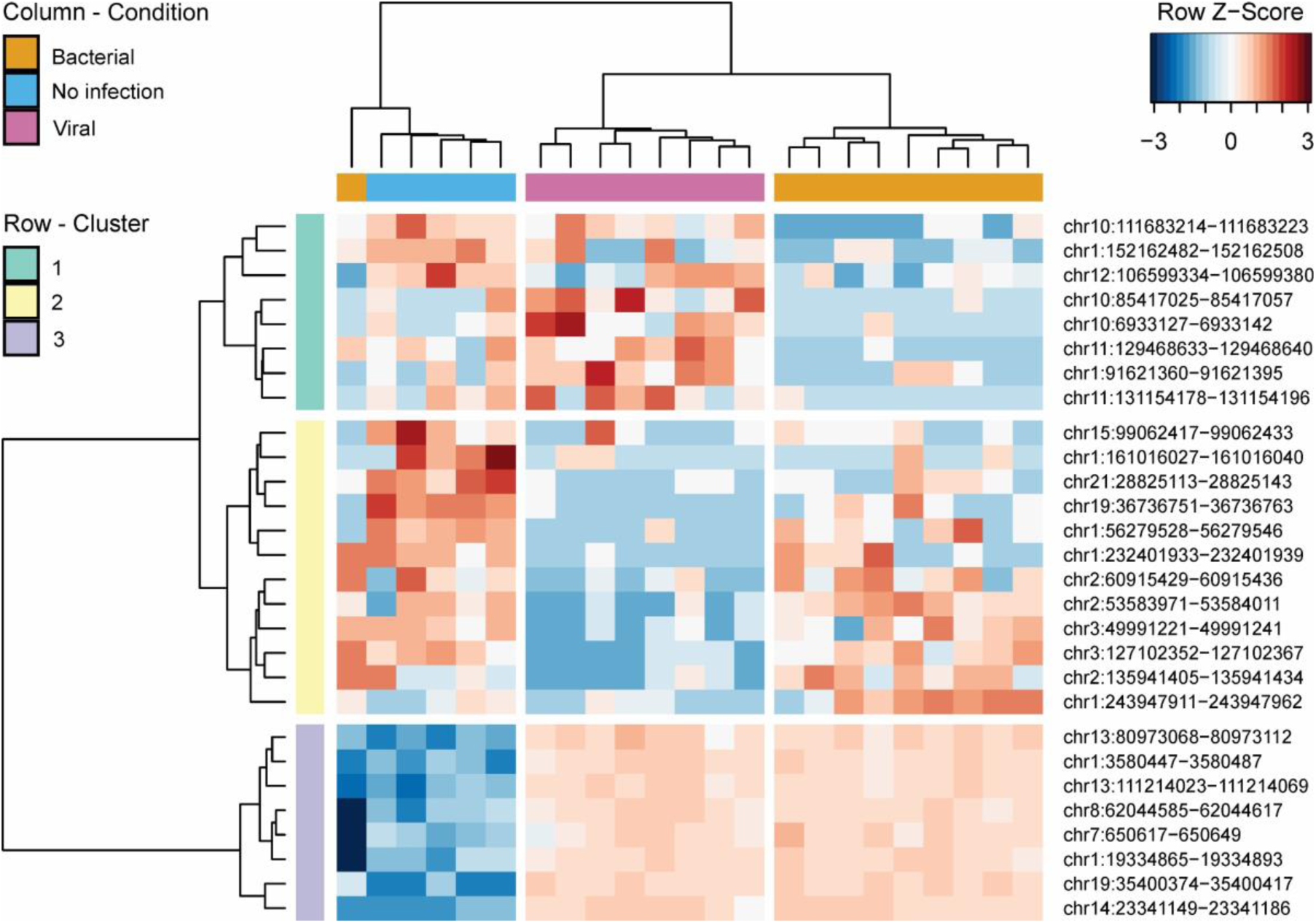
Heatmap and hierarchical clustering of differential enrichment analysis of the infection type comparison of non-infected controls (blue), viral (pink) and bacterial sepsis (yellow). Based on the top 5 DER per condition, hierarchical clustering was performed. DER signals are normalized per row according to Z-score and color-coded.

**Supplement Figure 13:**
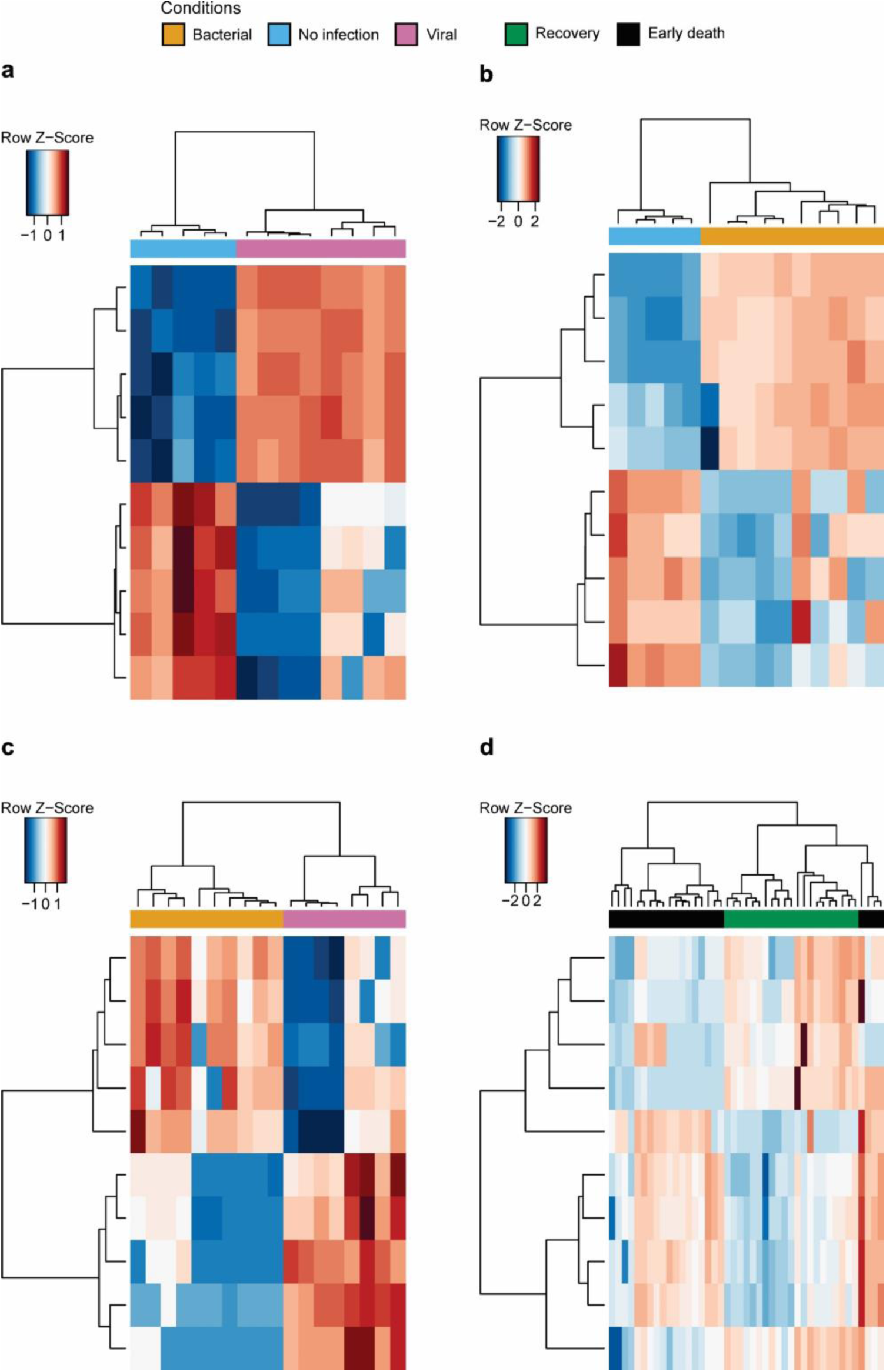
Heatmap and hierarchical clustering of differential enrichment analysis of the infection type and outcome cohort. Based on the top 5 DER per condition, hierarchical clustering was performed. DER signals are normalized per row according to Z-score and color-coded. a) Heatmap for DERs for bacterial (yellow) in comparison to viral sepsis (pink). b) Heatmap for DERs for bacterial sepsis (yellow) in comparison to non-infected, postoperative controls (blue). c) Heatmap for DERs for viral sepsis (pink) in comparison to non-infected, postoperative controls (blue). d) Heatmap for DERs for early death (black) in compaison to recovery in sepsis (green).

**Supplement Figure 14:**
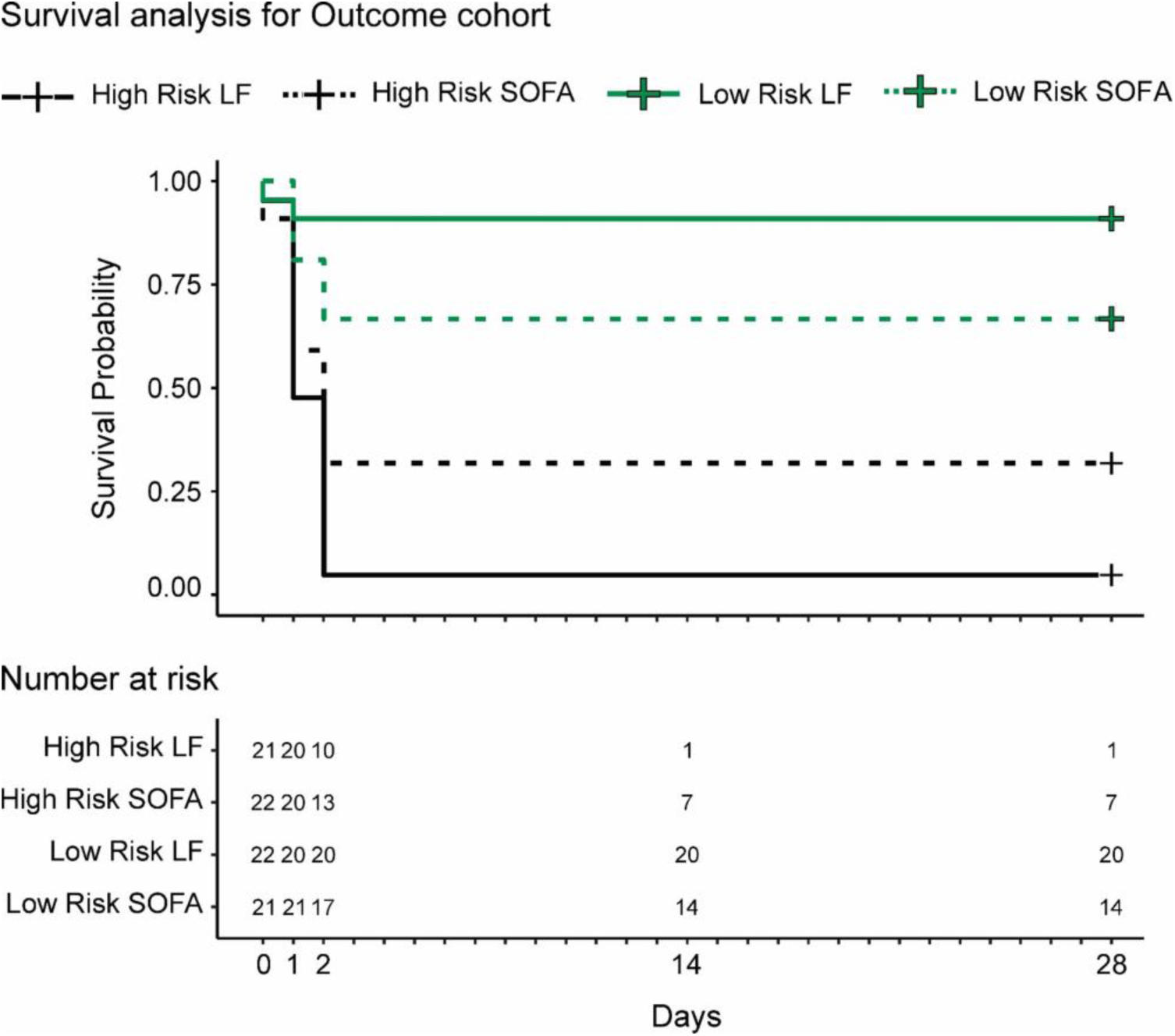
Time-to-event analysis comparing footprint DNA and SOFA risk groups. Based on disease severity scoring with SOFA and footprint DNA signals, 43 patients from the outcome cohort were assigned to the low risk SOFA (SOFA ≤ 6; green dashed line) or high risk SOFA (Sofa > 6; black dashed line) and to the low risk footprint DNA (low risk LF, green line) or the high risk footprint DNA group (high risk LF, black line). Over the course of 28 days, the survival probability is depicted for each group.

**Supplement Table 1:**
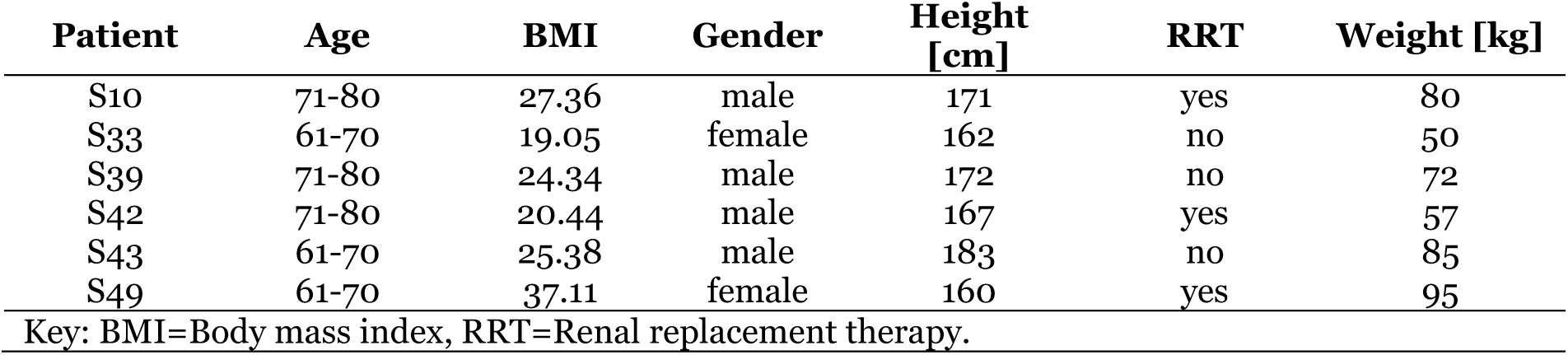
Patient metadata summary for sepsis cases over time (physiological status cohort).

**Supplement Table 2:** Patient metadata summary for sepsis cases over time for each physiological parameter and time point (physiological status cohort identification).

| Patient S10 |  |  |  |  |  |  |
| --- | --- | --- | --- | --- | --- | --- |
| Time point | To | T1 | T2 | T3 | T4 | T5 |
| Albumin [g/L] | 20.2 | 12.2 | 13.6 | 20.1 | 21.3 | NA |
| ALT [U/L] | 96 | 104 | 62 | 77 | 37 | NA |
| AP [U/L] | 73 | 60 | 78 | 96 | 389 | NA |
| aPTT [s] | 29.3 | 31.6 | 31.6 | 35.7 | 38.8 | NA |
| AST [U/L] | 40 | 139 | 304 | 92 | 135 | NA |
| Bilirubin [μM] | 10.2624 | 28.56368 | 13.6832 | 24.45872 | 51.312 | NA |
| Creatinine [μM] | 162.42 | 205.89 | 138.78 | 186.06 | 77.78 | NA |
| CRP [mg/dL] | 40.89 | 28.28 | 31.93 | 24.26 | 16.31 | NA |
| Erythrocytes [1/pL] | 2.9 | 2.4 | 2.3 | 2.7 | 2.4 | NA |
| Hemoglobine [g/dL] | 8.2 | 7 | 6.9 | 7.9 | 7.2 | NA |
| Hematocrite [%] | 25 | 21 | 20 | 24 | 21 | NA |
| Leukocytes [gpt/I] | 10.37 | 16.28 | 11.9 | 10.06 | 8.58 | NA |
| PCT [ng/mL] | 6.40 | 11.67 | 9.96 | 6.80 | 4.88 | NA |
| Potassium [mM] | 4.7 | 5.87 | 4.39 | 4.96 | 4.26 | NA |
| Quick [%] | 87.9 | 64.9 | 92.1 | 57.2 | 109 | NA |
| Sodium [mM] | 145 | 142 | 143 | 139 | 133 | NA |
| SOFA | 6 | 13 | 13 | 13 | 14 | NA |
| Thrombocytes [gpt/L] | 501 | 319 | 243 | 273 | 125 | NA |
| Urea [mM] | 15.984 | 17.316 | 10.323 | 12.8205 | 6.1605 | NA |

| Patient S33 |  |  |  |  |  |  |
| --- | --- | --- | --- | --- | --- | --- |
| Time point | To | T1 | T2 | T3 | T4 | T5 |
| Albumin [g/L] | 5.8 | 23.2 | 23.3 | 20 | 22.9 | 24.1 |
| ALT [U/L] | 43 | 18 | 18 | 6 | 9 | 8 |
| AP [U/L] | 35 | 46 | 122 | 125 | 192 | 172 |
| aPTT [s] | 56.5 | 60.1 | 42.8 | 25.8 | 25.2 | 24.8 |
| AST [U/L] | 93 | 39 | 34 | 14 | 17 | 12 |
| Bilirubin [μM] | 3.4208 | 18.8144 | 8.552 | 3.4208 | 3.4208 | 3.4208 |
| Creatinine [μM] | 43.46 | 59.48 | 48.80 | 44.23 | 36.60 | 34.31 |
| CRP [mg/dL] | 2.83 | 15.87 | 29.83 | 15.49 | 15.63 | 11.95 |
| Erythrocytes [1/pL] | 3.4 | 3.2 | 3.7 | 3 | 3 | 3.8 |
| Hemoglobine [g/dL] | 9.5 | 8.6 | 10.5 | 8.2 | 8.7 | 10.2 |
| Hematocrite [%] | 29 | 26 | 32 | 25 | 26 | 31 |
| Leukocytes [gpt/I] | 2.07 | 7.46 | 19.68 | 23.16 | 21.61 | 20.66 |
| PCT [ng/mL] | 55.91 | 30.32 | 12.63 | 0.53 | 0.16 | 0.11 |
| Potassium [mM] | 3.98 | 3.94 | NA | NA | NA | 4.35 |
| Quick [%] | 30.7 | 40.5 | 66 | 63.4 | 61.8 | 63.4 |
| Sodium [mM] | 149 | 146 | NA | NA | NA | 141 |
| SOFA | 11 | 11 | 11 | 11 | 8 | 8 |
| Thrombocytes [gpt/L] | 345 | 160 | 105 | 207 | 612 | 634 |
| Urea [mM] | 5.328 | 5.661 | 6.1605 | 10.989 | 5.4945 | 5.4945 |

| Patient S39 |  |  |  |  |  |  |
| --- | --- | --- | --- | --- | --- | --- |
| Time point | To | T1 | T2 | T3 | T4 | T5 |
| Albumin [g/L] | 19.9 | 24.9 | 22.3 | 27.4 | NA | NA |
| ALT [U/L] | 13 | 19 | 70 | 24 | NA | NA |
| AP [U/L] | 101 | 81 | 92 | 150 | NA | NA |
| aPTT [s] | 41.2 | 34.8 | 30.2 | 29 | NA | NA |
| AST [U/L] | 30 | 52 | 128 | 44 | NA | NA |
| Bilirubin [μM] | 99.2032 | 109.4656 | 106.0448 | 25.656 | NA | NA |
| Creatinine [μM] | 70.92 | 73.20 | 76.25 | 69.39 | NA | NA |
| CRP [mg/dL] | 21.66 | 20.76 | 13.45 | 11.65 | NA | NA |
| Erythrocytes [1/pL] | 2.5 | 2.5 | 2.5 | 2.2 | NA | NA |
| Hemoglobine [g/dL] | 7.4 | 7.9 | 8 | 6.7 | NA | NA |
| Hematocrite [%] | 23 | 23 | 23 | 21 | NA | NA |
| Leukocytes [gpt/I] | 16.74 | 9.33 | 4.77 | 3.46 | NA | NA |
| PCT [ng/mL] | 2.75 | 2.32 | 1.94 | 0.56 | NA | NA |
| Potassium [mM] | 4.73 | 4.81 | 4.37 | 4.15 | NA | NA |
| Quick [%] | 59.1 | 69.9 | 83.5 | 80.8 | NA | NA |
| Sodium [mM] | 138 | 143 | 147 | 139 | NA | NA |
| SOFA | 15 | 15 | 15 | 2 | NA | NA |
| Thrombocytes [gpt/L] | 92 | 69 | 78 | 197 | NA | NA |
| Urea [mM] | 6.66 | 7.8255 | 9.324 | 5.4945 | NA | NA |

| Patient S42 |  |  |  |  |  |  |
| --- | --- | --- | --- | --- | --- | --- |
| Time point | To | T2 | T3 | T4 | T5 | T5 |
| Albumin [g/L] | 31 | NA | NA | 12.1 | 26.3 | 30.6 |
| ALT [U/L] | 5 | NA | NA | 4 | 5 | 5 |
| AP [U/L] | 45 | NA | NA | 223 | 183 | 137 |
| aPTT [s] | 25.3 | NA | NA | 47.9 | 43.6 | 25.3 |
| AST [U/L] | 14 | NA | NA | 15 | 16 | 16 |
| Bilirubin [μM] | 6.8416 | NA | NA | 5.1312 | 3.4208 | 3.4208 |
| Creatinine [μM] | 93.79 | NA | NA | 136.49 | 92.27 | 77.78 |
| CRP [mg/dL] | 15.16 | NA | NA | 19.33 | 15.37 | 10.37 |
| Erythrocytes [1/pL] | 4 | NA | NA | 2.8 | 2.3 | 2.7 |
| Hemoglobine [g/dL] | 11.7 | NA | NA | 8.3 | 6.9 | 7.9 |
| Hematocrite [%] | 34 | NA | NA | 25 | 21 | 24 |
| Leukocytes [gpt/I] | 2.12 | NA | NA | 13.04 | 9.04 | 10.95 |
| PCT [ng/mL] | 32.90 | NA | NA | 2.87 | 0.78 | 0.19 |
| Potassium [mM] | 3.71 | NA | NA | 4.46 | 4.14 | 4.01 |
| Quick [%] | 91 | NA | NA | 91 | 110 | 101.3 |
| Sodium [mM] | 136 | NA | NA | 141 | 134 | 132 |
| SOFA | 14 | NA | NA | 13 | 8 | 6 |
| Thrombocytes [gpt/L] | 156 | NA | NA | 147 | 468 | 350 |
| Urea [mM] | 6.1605 | NA | NA | 10.656 | 5.994 | 6.1605 |

| Patient S43 |  |  |  |  |  |  |
| --- | --- | --- | --- | --- | --- | --- |
| Time point | To | T1 | T2 | T3 | T4 | T5 |
| Albumin [g/L] | 30.7 | 14.1 | 15.5 | 27.1 | NA | NA |
| ALT [U/L] | 22 | 23 | 18 | 29 | NA | NA |
| AP [U/L] | 36 | 76 | 57 | 90 | NA | NA |
| aPTT [s] | 50.2 | 37.8 | 38.7 | 27.2 | NA | NA |
| AST [U/L] | 35 | 40 | 27 | 40 | NA | NA |
| Bilirubin [μM] | 6.8416 | 6.8416 | 5.1312 | 6.8416 | NA | NA |
| Creatinine [μM] | 106.76 | 79.30 | 73.97 | 63.29 | NA | NA |
| CRP [mg/dL] | 43.03 | 49.99 | 43.01 | 18.11 | NA | NA |
| Erythrocytes [1/pL] | 2.1 | 2.6 | 2.2 | 2.4 | NA | NA |
| Hemoglobine [g/dL] | 7.7 | 7.9 | 7 | 7.4 | NA | NA |
| Hematocrite [%] | 22 | 25 | 22 | 22 | NA | NA |
| Leukocytes [gpt/I] | 2.15 | 12.41 | 11.31 | 13.55 | NA | NA |
| PCT [ng/mL] | 36.14 | 41.07 | 24.38 | 1.70 | NA | NA |
| Potassium [mM] | 4.17 | 4.23 | 4.09 | 3.95 | NA | NA |
| Quick [%] | 82.4 | 85.3 | 111 | 91 | NA | NA |
| Sodium [mM] | 145 | 150 | 152 | 141 | NA | NA |
| SOFA | 12 | 12 | 12 | 4 | NA | NA |
| Thrombocytes [gpt/L] | 187 | 128 | 152 | 165 | NA | NA |
| Urea [mM] | 8.8245 | 7.8255 | 9.4905 | 7.1595 | NA | NA |

| Patient S49 |  |  |  |  |  |  |
| --- | --- | --- | --- | --- | --- | --- |
| Time point | To | T1 | T2 | T3 | T4 | T5 |
| Albumin [g/L] | 24.1 | NA | 17.6 | NA | NA | NA |
| ALT [U/L] | 23 | NA | 19 | 24 | 22 | 17 |
| AP [U/L] | 72 | NA | 66 | 123 | 204 | 219 |
| aPTT [s] | 32.3 | NA | 40.1 | 25.9 | 29.1 | 24 |
| AST [U/L] | 63 | NA | 118 | 114 | 43 | 26 |
| Bilirubin [μM] | 3.4208 | NA | 11.9728 | 10.2624 | 5.1312 | 6.8416 |
| Creatinine [μM] | 275.28 | NA | 128.87 | 50.33 | 39.65 | 30.50 |
| CRP [mg/dL] | NA | NA | 29.45 | 12.07 | 18.67 | 5.11 |
| Erythrocytes [1/pL] | 3.5 | NA | 3 | 2.9 | 2.4 | 2.4 |
| Hemoglobine [g/dL] | 10.4 | NA | 9.4 | 8.7 | 7.3 | 7.5 |
| Hematocrite [%] | 29 | NA | 25 | 25 | 21 | 21 |
| Leukocytes [gpt/I] | 4.13 | NA | 3.62 | 1.07 | 1.23 | 5.15 |
| PCT [ng/mL] | 3.71 | NA | 2.02 | 0.30 | 0.14 | 0.05 |
| Potassium [mM] | 4.6 | NA | 4.77 | 4.18 | 4.2 | 3.8 |
| Quick [%] | 50.9 | NA | 92.8 | 69.9 | 96.9 | 89.2 |
| Sodium [mM] | 140 | NA | 141 | 161 | 150 | 140 |
| SOFA | 17 | NA | 14 | 8 | 4 | 1 |
| Thrombocytes [gpt/L] | 70 | NA | 62 | 32 | 53 | 341 |
| Urea [mM] | 41.625 | NA | 18.1485 | 21.312 | 11.322 | 7.659 |

**Supplement Table 3.**
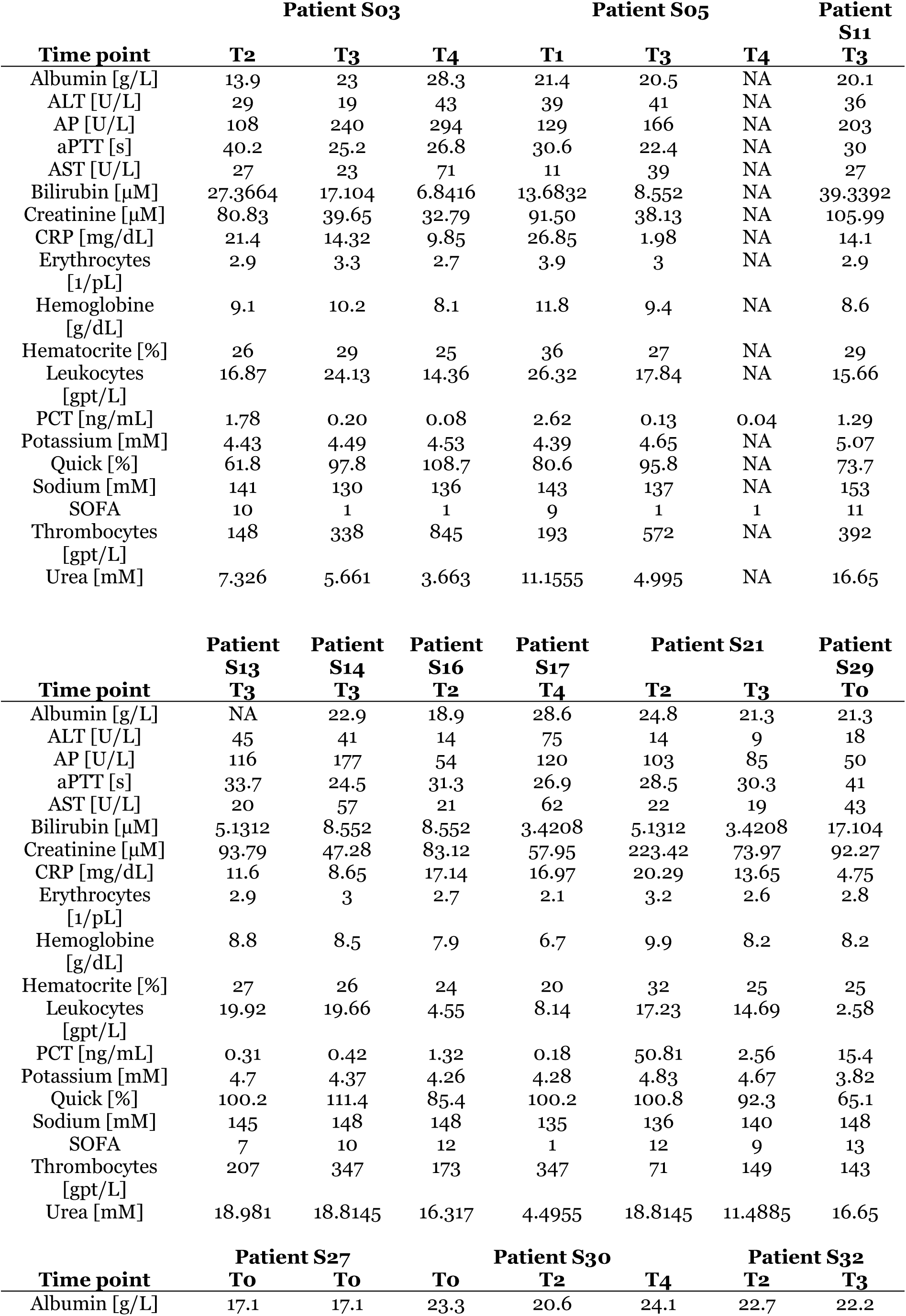
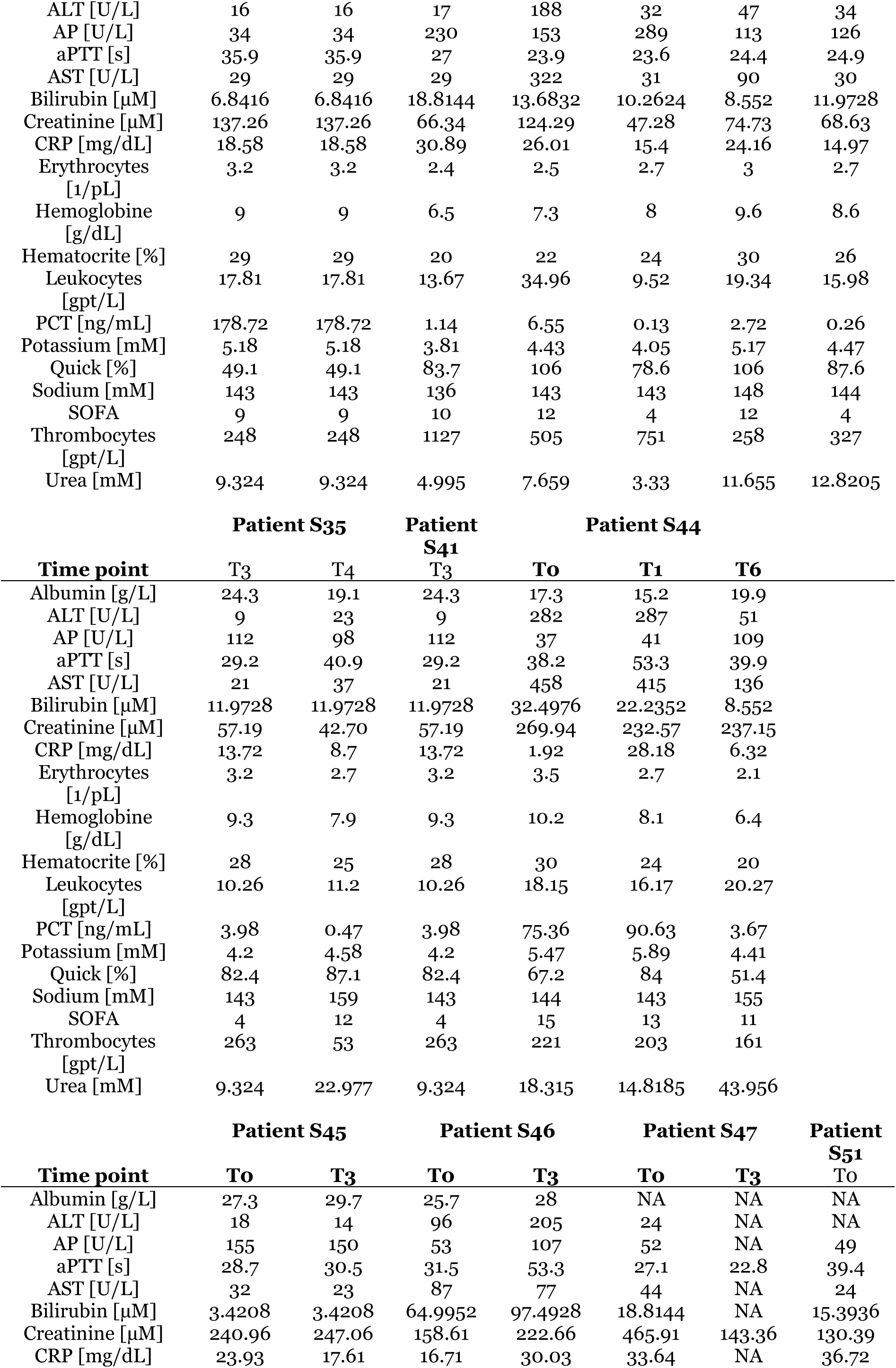
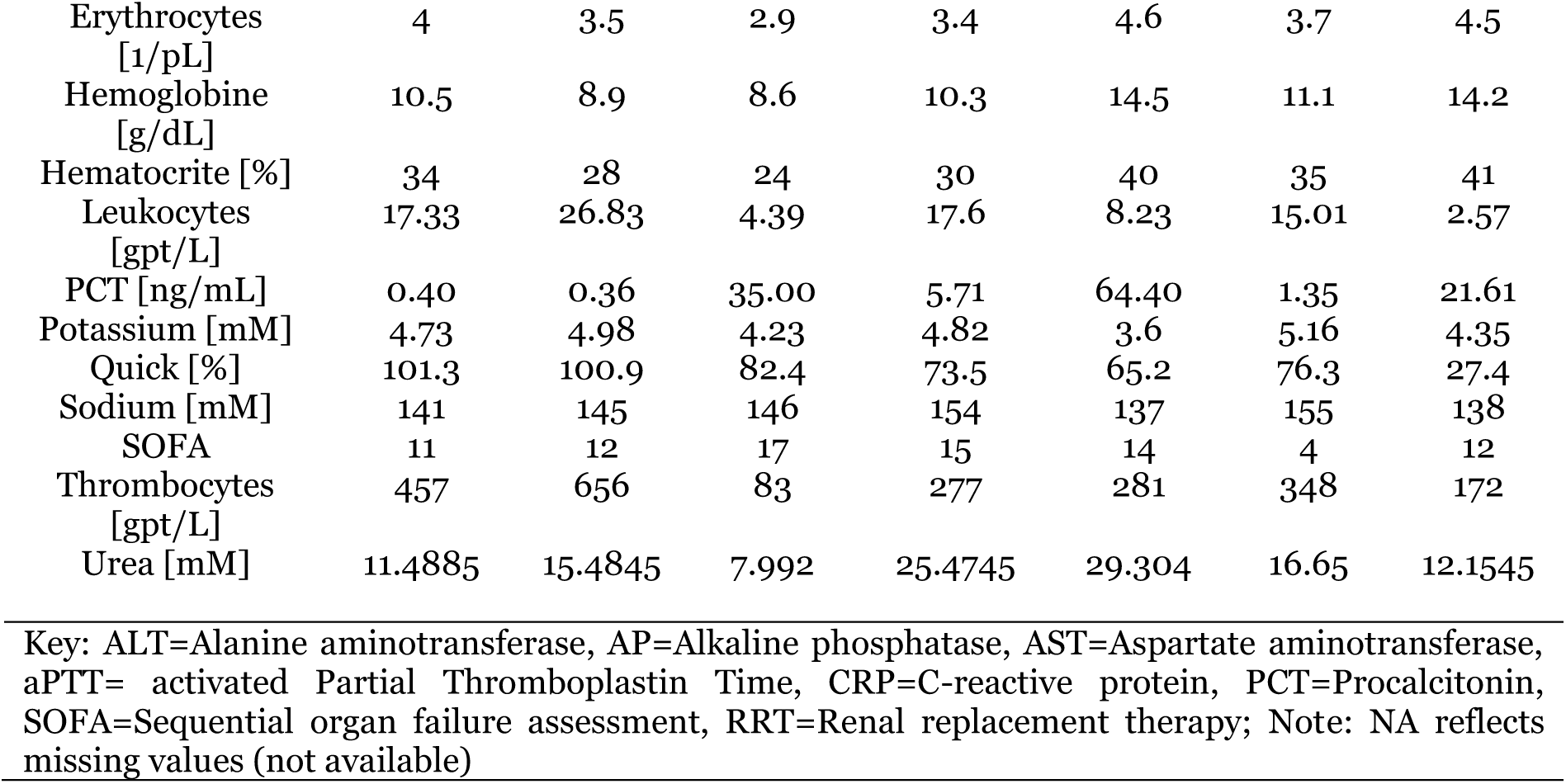
Patient metadata summary for sepsis cases over time for each physiological parameter and time point from the validation set (physiological status test validation cohort). NA=not available

**Supplement Table 4.**
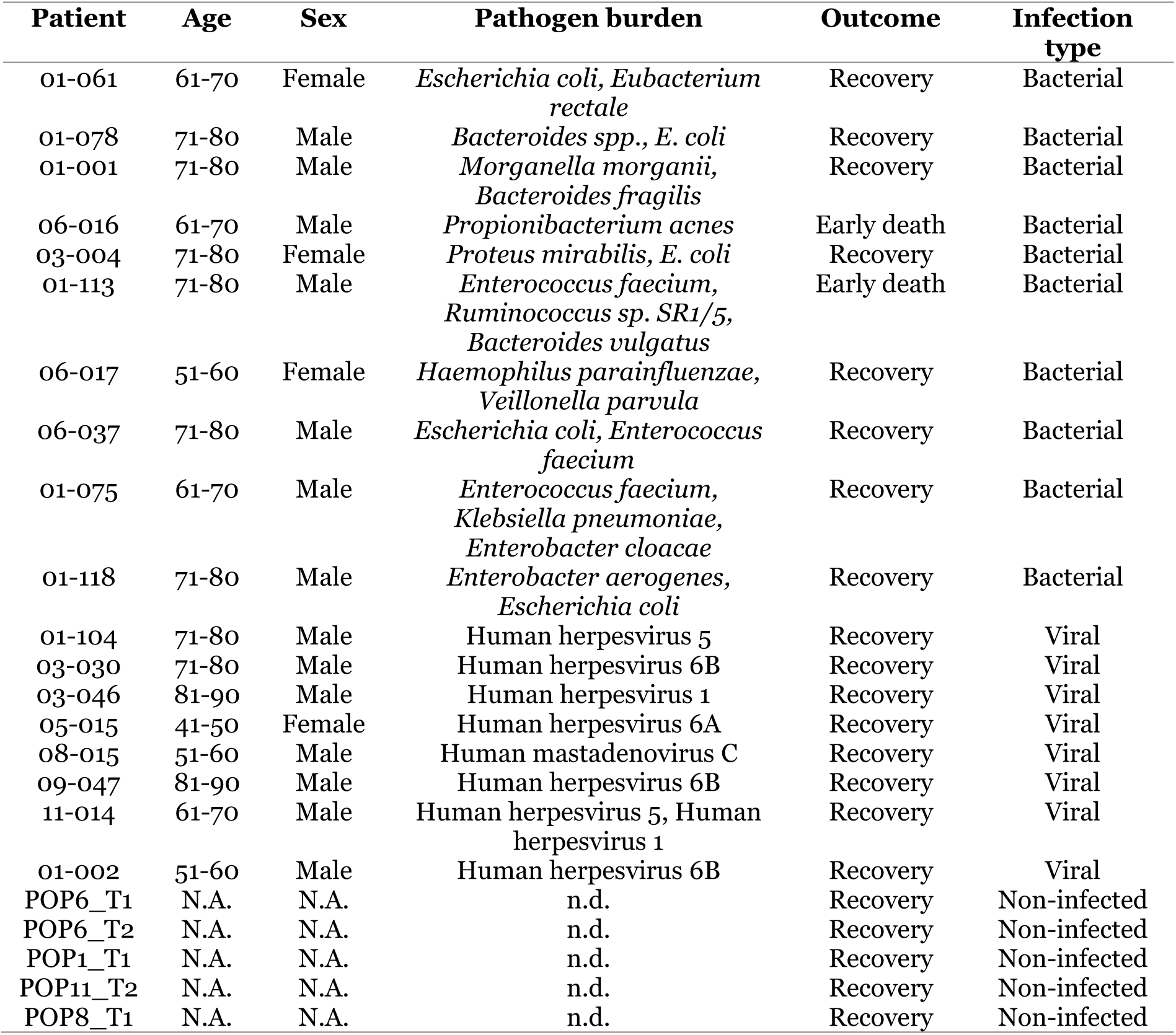
Patient descriptive data, pathogen burden, outcome and infection type summary for sepsis cases in infection type cohort. Early death reflects patients that died within two days after ICU admission. Survivors reached the end of the study at 28 days after ICU admission.

**Supplement Table 5.** Patient metadata summary statistics for bacterial and viral sepsis cases (infection type cohort). SD = Standard deviation, IQ = Inter quartile

| **Infection type** |  |  |
| --- | --- | --- |
|  | **Bacterial** | **Viral** |
| Age |  |  |
| N | 10 | 8 |
| Mean (SD) | 71.1 (6.79) | 66.4 (15.69) |
| Median | 71.5 | 69.5 |
| Range | (57; 79) | (41; 86) |
| IQ range | (68.0; 76.0) | (54.0; 78.5) |
| Albumin [g/L] |  |  |
| N | 9 | 6 |
| Mean (SD) | 27.73 (2.310) | 24.78 (7.776) |
| Median | 26.60 | 26.20 |
| Range | (25.7; 32.2) | (11.0; 32.5) |
| IQ range | (26.00; 29.00) | (21.75; 31.00) |
| ALT [U/L] |  |  |
| N | 10 | 7 |
| Mean (SD) | 148.1 (158.39) | 465.7 (734.08) |
| Median | 67.0 | 268.5 |
| Range | (22; 473) | (24; 2086) |
| IQ range | (43.0; 291.0) | (48.0; 494.0) |
| AP [U/L] |  |  |
| N | 10 | 6 |
| Mean (SD) | 120.1 (66.60) | 106.7 (62.46) |
| Median | 104.5 | 89.0 |
| Range | (63; 270) | (48; 192) |
| IQ range | (76.0; 122.5) | (48.6; 174.0) |
| aPTT [s] |  |  |
| N | 10 | 8 |
| Mean (SD) | 39.0 (10.08) | 34.9 (7.94) |
| Median | 38.5 | 33.8 |
| Range | (25; 58) | (24; 48) |
| IQ range | (33.0; 47.0) | (29.5; 40.5) |
| AST [U/L] |  |  |
| N | 10 | 6 |
| Mean (SD) | 240.6 (316.32) | 751.2 (648.67) |
| Median | 117.0 | 685.0 |
| Range | (34; 1080) | (72; 1747) |
| IQ range | (68.5; 294.0) | (128.0; 1190.0) |
| Bilirubin [μM] |  |  |
| N | 10 | 8 |
| Mean (SD) | 31.984 (24.2498) | 19.087 (16.5312) |
| Median | 28.222 | 10.981 |
| Range | (4.28; 80.39) | (8.55; 47.89) |
| IQ range | (12.828; 53.878) | (8.980; 28.164) |
| BMI |  |  |
| N | 10 | 8 |
| Mean (SD) | 26.12 (3.748) | 26.16 (4.787) |
| Median | 27.20 | 26.99 |
| Range | (17.9; 30.1) | (18.3; 31.8) |
| IQ range | (24.62; 29.07) | (22.33; 30.27) |
| Creatinine [μM] |  |  |
|  | **Bacterial** | **Viral** |
| N | 10 | 8 |
| Mean (SD) | 137.600 (91.0104) | 220.083 (139.6112) |
| Median | 88.836 | 179.197 |
| Range | (56.05; 269.94) | (61.00; 494.50) |
| IQ range | (73.204; 264.220) | (129.043; 294.340) |
| CRP [mg/dL] |  |  |
| N | 10 | 7 |
| Mean (SD) | 19.04 (12.300) | 19.47 (10.791) |
| Median | 17.25 | 15.53 |
| Range | (3.1; 39.4) | (5.8; 37.1) |
| IQ range | (12.17; 27.02) | (11.90; 29.03) |
| Sex |  |  |
| N | 10 | 8 |
| Male | 7 (70.0%) | 7 (87.5%) |
| Female | 3 (30.0%) | 1 (12.5%) |
| GFR [mL/min] |  |  |
| N | 9 | 8 |
| Mean (SD) | 53.28 (23.440) | 34.10 (22.929) |
| Median | 57.50 | 26.90 |
| Range | (16.4; 87.0) | (10.8; 81.0) |
| IQ range | (52.95; 59.70) | (18.28; 45.33) |
| Hematocrit [%] |  |  |
| N | 10 | 8 |
| Mean (SD) | 28.9 (4.13) | 30.9 (5.24) |
| Median | 27.9 | 30.1 |
| Range | (24; 38) | (25; 41) |
| IQ range | (26.0; 29.9) | (26.8; 33.5) |
| INR |  |  |
| N | 10 | 8 |
| Mean (SD) | 1.540 (0.3237) | 1.326 (0.4704) |
| Median | 1.450 | 1.253 |
| Range | (1.26; 2.37) | (0.90; 2.40) |
| IQ range | (1.305; 1.600) | (1.025; 1.375) |
| Lactate [mM] |  |  |
| N | 10 | 8 |
| Mean (SD) | 6.473 (5.3764) | 2.866 (1.9934) |
| Median | 4.249 | 2.077 |
| Range | (1.00; 18.97) | (0.65; 5.98) |
| IQ range | (3.258; 9.100) | (1.325; 4.750) |
| Leukocytes [gpt/I] |  |  |
| N | 10 | 8 |
| Mean (SD) | 16.018 (8.9481) | 16.486 (6.9768) |
| Median | 16.223 | 14.860 |
| Range | (2.17; 34.80) | (9.55; 30.73) |
| IQ range | (8.900; 20.400) | (11.085; 19.860) |
| PCT [ng/mL] |  |  |
| N | 10 | 8 |
| Mean (SD) | 55.01 (80.582) | 10.73 (11.803) |
| Median | 31.93 | 5.74 |
| Range | (2.1; 268.4) | (0.4; 34.3) |
| IQ range | (5.45; 55.96) | (2.14; 17.68) |
|  | **Bacterial** | **Viral** |
| Potassium [mM] |  |  |
| N | 10 | 8 |
| Mean (SD) | 4.75 (0.439) | 4.48 (0.542) |
| Median | 4.77 | 4.57 |
| Range | (4.0; 5.4) | (3.6; 5.2) |
| IQ range | (4.55; 4.95) | (4.08; 4.88) |
| Protein [g/L] |  |  |
| N | 7 | 5 |
| Mean (SD) | 45.37 (6.583) | 48.20 (9.549) |
| Median | 48.00 | 50.70 |
| Range | (32.5; 51.0) | (37.7; 59.6) |
| IQ range | (40.45; 49.10) | (39.00; 54.00) |
| Quick [%] |  |  |
| N | 10 | 8 |
| Mean (SD) | 50.0 (10.87) | 70.3 (27.90) |
| Median | 52.5 | 64.3 |
| Range | (28; 62) | (30; 111) |
| IQ range | (44.5; 58.0) | (53.0; 93.8) |
| RRT |  |  |
| N | 10 | 8 |
| No | 6 (60.0%) | 7 (87.5%) |
| Yes | 4 (40.0%) | 1 (12.5%) |
| Septic shock |  |  |
| N | 10 | 8 |
| No | 2 (20.0%) | 3 (37.5%) |
| Yes | 8 (80.0%) | 5 (62.5%) |
| Sodium [mM] |  |  |
| N | 10 | 8 |
| Mean (SD) | 144.65 (5.088) | 141.94 (8.287) |
| Median | 145.00 | 140.00 |
| Range | (136.5; 151.5) | (132.0; 155.0) |
| IQ range | (140.00; 148.00) | (135.50; 148.75) |
| SOFA |  |  |
| N | 10 | 7 |
| Mean (SD) | 9.7 (3.53) | 8.9 (3.13) |
| Median | 10.0 | 8.0 |
| Range | (4; 15) | (5; 13) |
| IQ range | (7.0; 12.0) | (6.0; 12.0) |
| Thrombocytes [gpt/I] |  |  |
| N | 10 | 8 |
| Mean (SD) | 194.7 (129.37) | 195.1 (141.16) |
| Median | 143.8 | 181.5 |
| Range | (42; 392) | (52; 494) |
| IQ range | (105.0; 330.0) | (85.5; 240.5) |
| Urea [mM] |  |  |
| N | 10 | 8 |
| Mean (SD) | 17.210 (12.5304) | 20.324 (6.1455) |
| Median | 12.446 | 20.754 |
| Range | (5.89; 43.00) | (9.74; 28.50) |
| IQ range | (8.242; 27.056) | (16.209; 25.213) |

Vasopressors or inotropes
| **Infection type** |  |  |
| --- | --- | --- |
|  | **Bacterial** | **Viral** |
| N | 10 | 8 |
| No | 1 (10.0%) | 1 (12.5%) |
| Yes | 9 (90.0%) | 7 (87.5%) |

**Supplement Table 6.** Patient metadata differences between bacterial and viral sepsis cases (infection type cohort). Categorical features are marked with an asterisk (‘*’). P values were obtained from two-sided Wilcoxon rank-sum test for numerical features, and Fisher’s exact test for categorical features. Area under ROC curves (‘AUROC’) is reported with 95% confidence interval boundaries (‘AUROC_ci_l’ = lower boundary, ‘AUROC_ci_h’ = higher boundary). From the ROC analysis per metric, the decision threshold (‘Thresh’) that maximizes the sum of specificity (‘Spec’) and sensitivity (‘Sens’) is indicated with the corresponding specificity and sensitivity. ALT=Alanine aminotransferase, AP=Alkaline phosphatase, AST=Aspartate aminotransferase, BMI=Body-Mass-Index, CRP=C-reactive protein, GFR=Glomerular filtration rate, INR=International normalized ratio, PCT=Procalcitonin, SOFA=Sequential organ failure assessment, RRT=Renal replacement therapy.

| **Infection type – Bacterial vs. Viral** |  |  |  |  |  |  |  |
| --- | --- | --- | --- | --- | --- | --- | --- |
| **Metric** | **P value** | **AUROC** | **AUROC_ci_l** | **AUROC_ci_h** | **Thresh** | **Spec** | **Sens** |
| Age | 0.689 | 0.563 | 0.236 | 0.889 | 64.500 | 0.900 | 0.500 |
| Albumin | 0.596 | 0.593 | 0.209 | 0.977 | 25.575 | 1.000 | 0.500 |
| ALT | 0.407 | 0.629 | 0.335 | 0.922 | 189.000 | 0.700 | 0.571 |
| AP | 0.447 | 0.625 | 0.299 | 0.951 | 103.000 | 0.600 | 0.667 |
| aPTT | 0.374 | 0.631 | 0.359 | 0.903 | 32.000 | 0.800 | 0.500 |
| AST | 0.093 | 0.767 | 0.497 | 1.000 | 446.750 | 0.900 | 0.667 |
| Bilirubin | 0.142 | 0.713 | 0.451 | 0.974 | 13.683 | 0.700 | 0.750 |
| BMI | 0.689 | 0.438 | 0.133 | 0.742 | 26.454 | 0.600 | 0.500 |
| Creatinine | 0.198 | 0.688 | 0.415 | 0.960 | 132.284 | 0.700 | 0.750 |
| CRP | 0.961 | 0.486 | 0.188 | 0.783 | 14.485 | 0.700 | 0.429 |
| Sex* | 0.588 | NA | NA | NA | NA | NA | NA |
| GFR | 0.112 | 0.736 | 0.460 | 1.000 | 51.800 | 0.778 | 0.875 |
| Hematocrit | 0.477 | 0.606 | 0.324 | 0.888 | 30.775 | 0.800 | 0.500 |
| INR | 0.061 | 0.769 | 0.511 | 1.000 | 1.375 | 0.700 | 0.750 |
| Lactate | 0.142 | 0.713 | 0.456 | 0.969 | 2.779 | 0.800 | 0.625 |
| Leukocytes | 1.000 | 0.500 | 0.211 | 0.789 | 15.890 | 0.600 | 0.625 |
| PCT | 0.083 | 0.750 | 0.515 | 0.985 | 8.068 | 0.700 | 0.625 |
| Potassium | 0.424 | 0.619 | 0.338 | 0.899 | 4.425 | 0.800 | 0.500 |
| Protein | 0.516 | 0.629 | 0.215 | 1.000 | 49.900 | 0.857 | 0.600 |
| Quick | 0.068 | 0.763 | 0.503 | 1.000 | 63.000 | 1.000 | 0.625 |
| RRT* | 0.314 | NA | NA | NA | NA | NA | NA |
| Septic_shock* | 0.608 | NA | NA | NA | NA | NA | NA |
| Sodium | 0.477 | 0.606 | 0.312 | 0.901 | 142.500 | 0.700 | 0.625 |
| SOFA | 0.659 | 0.571 | 0.280 | 0.863 | 8.500 | 0.600 | 0.571 |
| Thrombocytes | 0.894 | 0.525 | 0.232 | 0.818 | 151.500 | 0.600 | 0.625 |
| Urea | 0.230 | 0.675 | 0.390 | 0.960 | 15.751 | 0.700 | 0.875 |
| Vasopressors_inotropes* | 1.000 | NA | NA | NA | NA | NA | NA |

**Supplement Table 7.** Patient descriptive data, pathogen burden, infection type and outcome for sepsis cases in outcome cohort. Early death reflects septic patients that died within two days after ICU admission. Recovery reached the end of the study at 28 days after ICU admission.

| Patient | Age | Sex | Pathogen burden | Infection type | Outcome |
| --- | --- | --- | --- | --- | --- |
| 01-001 | 71-80 | Male | *Morganella morganii*,<br>*Bacteroides fragilis* | Bacterial | Early death |
| 01-002 | 51-60 | Male | Human herpesvirus 6B | Viral | Early death |
| 01-005 | 81-90 | Male | *Enterobacter cloacae*,<br>*Escherichia coli*, *Cronobacter sakazakii* | Bacterial | Early death |
| 01-025 | 71-80 | Female | *Escherichia coli*, *Klebsiella pneumoniae* | Bacterial | Early death |
| 01-036 | 71-80 | Male | *Enterobacter cloacae* | Bacterial | Early death |
| 01-049 | 81-90 | Male | *Escherichia coli* | Bacterial | Early death |
| 01-061 | 61-70 | Female | *Escherichia coli*, *Eubacterium rectale* | Bacterial | Early death |
| 01-075 | 61-70 | Male | *Enterococcus faecium*, *Klebsiella pneumoniae*, *Enterobacter cloacae* | Bacterial | Early death |
| 01-078 | 71-80 | Male | *Bacteroides* spp., *Escherichia coli* | Bacterial | Early death |
| 02-037 | 51-60 | Female | *Escherichia coli*, *Bacteroides fragilis*, *Bacteroides vulgatus*,<br>*Alistipes shahii* | Bacterial | Early death |
| 03-004 | 71-80 | Female | *Proteus mirabilis*, *Escherichia coli* | Bacterial | Early death |
| 03-009 | 51-50 | Male | *Escherichia coli*, *Bacteroides fragilis* | Bacterial | Early death |
| 06-017 | 51-60 | Female | *Haemophilus parainfluenzae*,<br>*Veillonella parvula* | Bacterial | Early death |
| 06-037 | 71-80 | Male | *Escherichia coli*, *Enterococcus faecium* | Bacterial | Early death |
| 08-005 | 71-80 | Male | *Enterobacter cloacae* | Bacterial | Early death |
| 08-015 | 51-60 | Male | Human mastadenovirus C | Viral | Early death |
| 08-026 | 71-80 | Male | *Escherichia coli* | Bacterial | Early death |
| 08-056 | 71-80 | Male | *Enterococcus faecalis* | Bacterial | Early death |
| 08-059 | 81-90 | Male | *Escherichia coli* | Bacterial | Early death |
| 09-031 | 71-80 | Male | *Escherichia coli* | Bacterial | Early death |
| 16-004 | 51-60 | Female | *Escherichia coli* | Bacterial | Early death |
| 01-006 | 71-80 | Male | *Staphylococcus aureus*, Torque<br>teno virus | Bacterial, Viral | Early death |
| 01-012 | 61-70 | Female | *Pseudomonas aeruginosa*,<br>*Lactobacillus fermentum* | Bacterial | Recovery |
| 01-064 | 51-60 | Female | *Escherichia coli* | Bacterial | Recovery |
| 01-070 | 71-80 | Female | *Streptococcus thermophilus*,<br>*Propionibacterium freudenreichii* | Bacterial | Recovery |
| 01-071 | 81-90 | Male | *Enterococcus faecium*, *Candida glabrata*, *Escherichia coli* | Bacterial,<br>Fungal | Recovery |
| 01-074 | 61-70 | Female | *Candida glabrata*, *Klebsiella oxytoca* | Bacterial,<br>Fungal | Recovery |
| 01-077 | 81-90 | Male | *Klebsiella oxytoca*, *Streptococcus pneumoniae*, *Lactobacillus salivarius*, Human herpesvirus<br>6B | Bacterial, Viral | Recovery |
| 01-086 | 81-90 | Female | *Staphylococcus aureus* | Bacterial | Recovery |
| 01-090 | 61-70 | Female | *Escherichia coli* | Bacterial | Recovery |
| 01-092 | 61-70 | Male | *Escherichia coli* | Bacterial | Recovery |
| 01-113 | 71-80 | Male | *Enterococcus faecium*,<br>*Ruminococcus* sp. SR1/5,<br>*Bacteroides vulgatus* | Bacterial | Recovery |
| 02-018 | 81-90 | Male | *Proteus mirabilis* | Bacterial | Recovery |
| 02-043 | 81-90 | Male | *Escherichia coli* | Bacterial | Recovery |
| 03-029 | 81-90 | Male | *Escherichia coli* | Bacterial | Recovery |
| 03-047 | 51-60 | Female | *Pseudomonas aeruginosa*,<br>*Escherichia coli* | Bacterial | Recovery |
| 04-004 | 41-50 | Male | *Clostridium perfringens* | Bacterial | Recovery |
| 05-005 | 51-60 | Male | *Roseburia intestinalis*,<br>*Escherichia coli*, *Bacteroides*<br>*vulgatus* | Bacterial | Recovery |
| 06-003 | 61-70 | Male | *Staphylococcus aureus* | Bacterial | Recovery |
| 06-016 | 61-70 | Male | *Propionibacterium acnes* | Bacterial | Recovery |
| 08-034 | 61-70 | Male | *Streptococcus spp.* | Bacterial | Recovery |
| 08-044 | 81-90 | Female | *Staphylococcus aureus*,<br>*Lactobacillus fermentum* | Bacterial | Recovery |
| 22-001 | 71-80 | Male | *Escherichia coli*, *Bacteroides spp.* | Bacterial | Recovery |

**Supplement Table 8.** Patient metadata summary statistics for early death and recovery sepsis cases (outcome cohort). SD = Standard deviation, IQ = Inter quartile.

| **Outcome** |  | **Early death** | **Recovery** |
| --- | --- | --- | --- |
| Age |  |  |  |
| N |  | 21 | 22 |
| Mean (SD) |  | 69.9 (10.76) | 70.5 (11.73) |
| Median |  | 74.0 | 70.5 |
| Range |  | (50; 84) | (45; 87) |
| IQ range |  | (57.0; 77.0) | (65.0; 82.0) |
| Albumin [g/L] |  |  |  |
| N |  | 15 | 19 |
| Mean (SD) |  | 27.61 (3.414) | 27.43 (5.033) |
| Median |  | 26.60 | 26.20 |
| Range |  | (21.8; 33.0) | (19.3; 39.0) |
| IQ range |  | (25.65; 31.10) | (23.00; 30.70) |
| ALT [U/L] |  |  |  |
| N |  | 16 | 19 |
| Mean (SD) |  | 87.3 (106.37) | 98.8 (145.46) |
| Median |  | 44.5 | 38.0 |
| Range |  | (7; 333) | (5; 509) |
| IQ range |  | (26.0; 71.0) | (21.0; 112.0) |
| AP [U/L] |  |  |  |
| N |  | 15 | 18 |
| Mean (SD) |  | 100.9 (52.90) | 132.2 (94.21) |
| Median |  | 82.0 | 114.3 |
| Range |  | (35; 206) | (23; 394) |
| IQ range |  | (60.0; 143.0) | (63.0; 194.0) |
| aPTT [s] |  |  |  |
| N |  | 21 | 22 |
| Mean (SD) |  | 33.1 (8.98) | 42.9 (13.90) |
| Median |  | 32.0 | 41.3 |
| Range |  | (24; 58) | (24; 79) |
| IQ range |  | (26.5; 36.5) | (32.0; 49.5) |
| AST [U/L] |  |  |  |
| N |  | 15 | 17 |
| Mean (SD) |  | 156.1 (295.78) | 216.4 (305.26) |
| Median |  | 63.0 | 72.0 |
| Range |  | (8; 1190) | (20; 1080) |
| IQ range |  | (33.5; 149.0) | (37.5; 177.0) |
| Bilirubin [μM] |  |  |  |
| N |  | 18 | 21 |
| Mean (SD) |  | 18.280 (12.9571) | 19.542 (13.9339) |
| Median |  | 14.966 | 17.000 |
| Range |  | (4.28; 54.73) | (3.42; 53.88) |
| IQ range |  | (10.262; 25.656) | (10.262; 22.235) |
| BMI |  |  |  |
| N |  | 21 | 21 |
| Mean (SD) |  | 27.62 (7.028) | 27.86 (10.385) |
| Median |  | 27.43 | 25.39 |
| Range |  | (17.9; 51.2) | (15.9; 69.2) |
| IQ range |  | (23.88; 29.38) | (23.88; 27.78) |
| Creatinine [μM] |  |  |  |

| Outcome |  |  |
| --- | --- | --- |
|  | Early death | Recovery |
| N | 21 | 22 |
| Mean (SD) | 155.931 (130.4415) | 199.389 (110.8686) |
| Median | 90.361 | 191.969 |
| Range | (32.79; 494.50) | (30.50; 513.00) |
| IQ range | (73.204; 181.485) | (136.495; 269.558) |
| CRP [mg/dL] |  |  |
| N | 20 | 20 |
| Mean (SD) | 18.48 (12.349) | 19.17 (10.833) |
| Median | 15.36 | 15.98 |
| Range | (3.1; 54.2) | (4.7; 44.9) |
| IQ range | (12.31; 25.95) | (12.24; 23.42) |
| Sex |  |  |
| N | 21 | 22 |
| Male | 15 (71.4%) | 14 (63.6%) |
| Female | 6 (28.6%) | 8 (36.4%) |
| GFR [mL/min] |  |  |
| N | 20 | 19 |
| Mean (SD) | 52.44 (30.011) | 28.22 (16.158) |
| Median | 55.50 | 22.50 |
| Range | (10.5; 101.9) | (9.1; 60.0) |
| IQ range | (21.58; 76.75) | (16.35; 39.65) |
| Hematocrit [%] |  |  |
| N | 21 | 22 |
| Mean (SD) | 29.9 (6.25) | 30.7 (4.75) |
| Median | 28.5 | 30.0 |
| Range | (19; 41) | (23; 38) |
| IQ range | (26.0; 36.0) | (25.9; 35.0) |
| INR |  |  |
| N | 21 | 22 |
| Mean (SD) | 1.394 (0.3392) | 1.681 (0.5847) |
| Median | 1.305 | 1.523 |
| Range | (0.90; 2.26) | (1.03; 3.70) |
| IQ range | (1.150; 1.500) | (1.330; 1.900) |
| Lactate [mM] |  |  |
| N | 21 | 22 |
| Mean (SD) | 4.795 (4.5987) | 7.386 (5.2218) |
| Median | 2.800 | 6.368 |
| Range | (0.75; 18.97) | (1.00; 18.70) |
| IQ range | (1.277; 6.000) | (2.581; 12.300) |
| Leukocytes [gpt/I] |  |  |
| N | 21 | 22 |
| Mean (SD) | 17.791 (10.1078) | 14.615 (12.6624) |
| Median | 15.820 | 10.550 |
| Range | (4.44; 43.68) | (0.66; 43.62) |
| IQ range | (10.320; 22.270) | (5.575; 23.500) |
| PCT [ng/mL] |  |  |
| N | 21 | 22 |
| Mean (SD) | 52.22 (86.922) | 65.10 (89.933) |
| Median | 5.87 | 14.39 |
| Range | (0.1; 268.4) | (0.6; 279.2) |
| IQ range | (2.14; 50.36) | (7.12; 100.00) |

| Outcome |  | Early death | Recovery |
| --- | --- | --- | --- |
| Potassium [mM] |  |  |  |
| N |  | 21 | 22 |
| Mean (SD) |  | 4.36 (0.703) | 4.67 (0.480) |
| Median |  | 4.35 | 4.71 |
| Range |  | (3.3; 5.4) | (3.5; 5.4) |
| IQ range |  | (3.95; 4.93) | (4.45; 5.00) |
| Protein [g/L] |  |  |  |
| N |  | 14 | 13 |
| Mean (SD) |  | 48.58 (8.706) | 44.80 (9.309) |
| Median |  | 49.20 | 43.70 |
| Range |  | (32.5; 60.1) | (29.7; 58.0) |
| IQ range |  | (40.45; 55.70) | (36.90; 55.00) |
| Quick [%] |  |  |  |
| N |  | 21 | 22 |
| Mean (SD) |  | 61.2 (20.57) | 51.9 (17.14) |
| Median |  | 58.0 | 54.3 |
| Range |  | (31; 111) | (15; 96) |
| IQ range |  | (49.5; 77.0) | (37.0; 60.0) |
| RRT |  |  |  |
| N |  | 21 | 22 |
| No |  | 18 (85.7%) | 16 (72.7%) |
| Yes |  | 3 (14.3%) | 6 (27.3%) |
| Septic shock |  |  |  |
| N |  | 21 | 22 |
| No |  | 7 (33.3%) | 2 (9.1%) |
| Yes |  | 14 (66.7%) | 20 (90.9%) |
| Sodium [mM] |  |  |  |
| N |  | 21 | 22 |
| Mean (SD) |  | 143.40 (5.183) | 140.41 (6.607) |
| Median |  | 144.00 | 138.75 |
| Range |  | (132.0; 151.5) | (128.0; 160.5) |
| IQ range |  | (140.00; 147.00) | (136.00; 143.50) |
| SOFA |  |  |  |
| N |  | 21 | 22 |
| Mean (SD) |  | 7.9 (3.68) | 11.4 (3.11) |
| Median |  | 8.0 | 11.0 |
| Range |  | (1; 14) | (6; 18) |
| IQ range |  | (5.0; 11.0) | (9.0; 13.0) |
| Thrombocytes [gpt/I] |  |  |  |
| N |  | 21 | 22 |
| Mean (SD) |  | 250.6 (143.36) | 161.0 (136.67) |
| Median |  | 240.0 | 110.0 |
| Range |  | (86; 697) | (40; 566) |
| IQ range |  | (140.0; 330.0) | (82.0; 193.5) |
| Urea [mM] |  |  |  |
| N |  | 21 | 22 |
| Mean (SD) |  | 14.119 (9.8640) | 22.721 (15.3671) |
| Median |  | 9.990 | 21.006 |
| Range |  | (5.00; 43.00) | (3.41; 55.85) |
| IQ range |  | (7.243; 18.200) | (11.405; 30.137) |
| Vasopressors or inotropes |  |  |  |

| **Outcome** |  |  |
| --- | --- | --- |
|  | **Early death** | **Recovery** |
| N | 21 | 22 |
| No | 6 (28.6%) | 1 (4.5%) |
| Yes | 15 (71.4%) | 21 (95.5%) |

**Supplement Table 9.** Patient metadata differences between early death and survival sepsis cases (outcome cohort). Categorical features are marked with an asterisk (‘*’). P values were obtained from two-sided Wilcoxon rank-sum test for numerical features, and Fisher’s exact test for categorical features. Metrics with a p value <= 0.05 are highlighted in bold font. Area under ROC curves (‘AUROC’) is reported with 95% confidence interval boundaries (‘AUROC_ci_l’ = lower boundary, ‘AUROC_ci_h’ = higher boundary). From the ROC analysis per metric, the decision threshold (‘Thresh’) that maximizes the sum of specificity (‘Spec’) and sensitivity (‘Sens’) is indicated with the corresponding specificity and sensitivity. ALT=Alanine aminotransferase, AP=Alkaline phosphatase, AST=Aspartate aminotransferase, BMI=Body-Mass-Index, CRP=C-reactive protein, GFR=Glomerular filtration rate, INR=International normalized ratio, PCT=Procalcitonin, SOFA=Sequential organ failure assessment, RRT=Renal replacement therapy.

| **Outcome – Early death vs. Recovery** |  |  |  |  |  |  |  |
| --- | --- | --- | --- | --- | --- | --- | --- |
| **Metric** | **P value** | **AUROC** | **AUROC_ci_l** | **AUROC_ci_h** | **Thresh** | **Spec** | **Sens** |
| Age | 0.990 | 0.498 | 0.318 | 0.678 | 71.500 | 0.571 | 0.591 |
| Albumin | 0.835 | 0.523 | 0.323 | 0.723 | 25.525 | 0.800 | 0.368 |
| ALT | 0.716 | 0.538 | 0.340 | 0.736 | 42.000 | 0.563 | 0.579 |
| AP | 0.459 | 0.578 | 0.376 | 0.779 | 106.500 | 0.733 | 0.556 |
| **aPTT** | **0.011** | **0.727** | **0.574** | **0.881** | **37.250** | **0.810** | **0.636** |
| AST | 0.571 | 0.561 | 0.355 | 0.767 | 70.250 | 0.600 | 0.529 |
| Bilirubin | 0.877 | 0.516 | 0.328 | 0.704 | 16.650 | 0.611 | 0.524 |
| BMI | 0.753 | 0.529 | 0.349 | 0.710 | 26.398 | 0.571 | 0.619 |
| Creatinine | 0.091 | 0.652 | 0.474 | 0.829 | 125.747 | 0.667 | 0.773 |
| CRP | 0.797 | 0.525 | 0.341 | 0.709 | 15.605 | 0.550 | 0.550 |
| Sex* | 0.747 | NA | NA | NA | NA | NA | NA |
| **GFR** | **0.018** | **0.724** | **0.556** | **0.892** | **47.275** | **0.650** | **0.842** |
| Hematocrit | 0.601 | 0.548 | 0.368 | 0.727 | 29.175 | 0.571 | 0.636 |
| **INR** | **0.039** | **0.685** | **0.523** | **0.847** | **1.323** | **0.571** | **0.773** |
| Lactate | 0.050 | 0.675 | 0.511 | 0.839 | 6.050 | 0.762 | 0.545 |
| Leukocytes | 0.178 | 0.621 | 0.447 | 0.795 | 13.485 | 0.667 | 0.591 |
| PCT | 0.220 | 0.610 | 0.434 | 0.786 | 9.780 | 0.619 | 0.682 |
| Potassium | 0.138 | 0.633 | 0.459 | 0.808 | 4.398 | 0.524 | 0.773 |
| Protein | 0.244 | 0.635 | 0.416 | 0.853 | 44.900 | 0.714 | 0.615 |
| Quick | 0.198 | 0.616 | 0.445 | 0.787 | 58.500 | 0.476 | 0.727 |
| RRT* | 0.457 | NA | NA | NA | NA | NA | NA |
| Septic_shock* | 0.069 | NA | NA | NA | NA | NA | NA |
| **Sodium** | **0.030** | **0.694** | **0.531** | **0.857** | **139.250** | **0.810** | **0.545** |
| **SOFA** | **0.003** | **0.760** | **0.617** | **0.902** | **9.500** | **0.667** | **0.682** |
| **Thrombocytes** | **0.008** | **0.738** | **0.585** | **0.891** | **118.250** | **0.857** | **0.591** |
| **Urea** | **0.048** | **0.677** | **0.510** | **0.845** | **11.280** | **0.571** | **0.773** |
| **Vasopressors_inotropes\*** | **0.046** | **NA** | **NA** | **NA** | **NA** | **NA** | **NA** |

