## Supplementary material for "Human *in vivo* footprints from blood plasma samples for improved diagnostics in septic patients": Publication licenses: Publication License Jan-10-2025_Fig3a.pdf

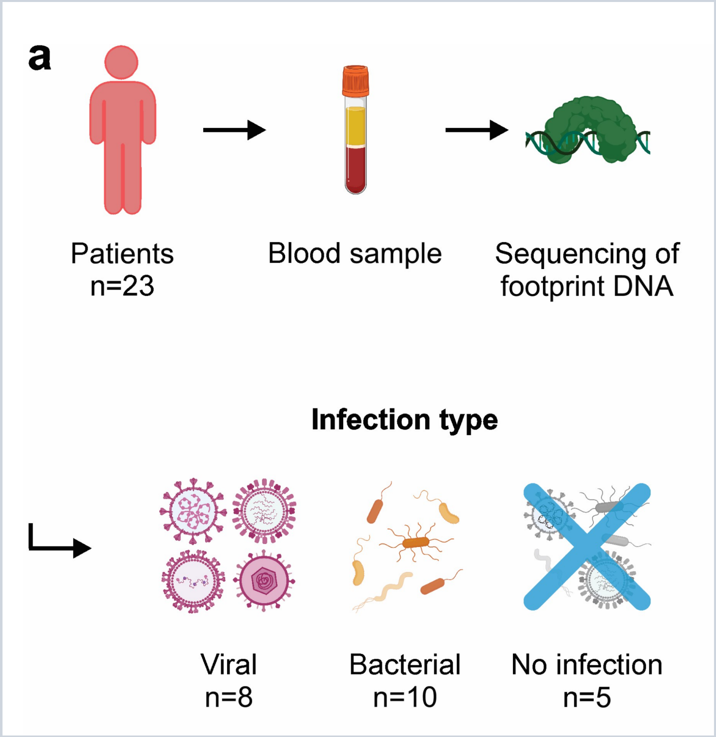


*For any questions regarding this document, or other questions about publishing with BioRender, please refer to our BioRender Publication Guide, or contact BioRender Support at.*
